# Impact of a blood-stage vaccine on *Plasmodium vivax* malaria

**DOI:** 10.1101/2022.05.27.22275375

**Authors:** Mimi M. Hou, Jordan R. Barrett, Yrene Themistocleous, Thomas A. Rawlinson, Ababacar Diouf, Francisco J. Martinez, Carolyn M. Nielsen, Amelia M. Lias, Lloyd D. W. King, Nick J. Edwards, Nicola M. Greenwood, Lucy Kingham, Ian D. Poulton, Baktash Khozoee, Cyndi Goh, Dylan J. Mac Lochlainn, Jo Salkeld, Micheline Guilotte-Blisnick, Christèle Huon, Franziska Mohring, Jenny M. Reimer, Virander S. Chauhan, Paushali Mukherjee, Sumi Biswas, Iona J. Taylor, Alison M. Lawrie, Jee-Sun Cho, Fay L. Nugent, Carole A. Long, Robert W. Moon, Kazutoyo Miura, Sarah E. Silk, Chetan E. Chitnis, Angela M. Minassian, Simon J. Draper

## Abstract

**Background:** There are no licensed vaccines against *Plasmodium vivax*, the most common cause of malaria outside of Africa.

**Methods:** We conducted two Phase I/IIa clinical trials to assess the safety, immunogenicity and efficacy of two vaccines targeting region II of *P. vivax* Duffy-binding protein (PvDBPII). Recombinant viral vaccines (using ChAd63 and MVA vectors) were administered at 0, 2 months or in a delayed dosing regimen (0, 17, 19 months), whilst a protein/adjuvant formulation (PvDBPII/Matrix-M™) was administered monthly (0, 1, 2 months) or in a delayed dosing regimen (0, 1, 14 months). Delayed regimens were due to trial halts during the COVID-19 pandemic. Volunteers underwent heterologous controlled human malaria infection (CHMI) with blood-stage *P. vivax* parasites at 2-4 weeks following their last vaccination, alongside unvaccinated controls. Efficacy was assessed by comparison of parasite multiplication rate (PMR) in blood post-CHMI, modelled from parasitemia measured by quantitative polymerase-chain-reaction (qPCR).

**Results:** Thirty-two volunteers were enrolled and vaccinated (n=16 for each vaccine). No safety concerns were identified. PvDBPII/Matrix-M™, given in the delayed dosing regimen, elicited the highest antibody responses and reduced the mean PMR following CHMI by 51% (range 36-66%; n=6) compared to unvaccinated controls (n=13). No other vaccine or regimen impacted parasite growth. *In vivo* growth inhibition of blood-stage *P. vivax* correlated with functional antibody readouts of vaccine immunogenicity.

**Conclusions:** Vaccination of malaria-naïve adults with a delayed booster regimen of PvDBPII/ Matrix-M™ significantly reduces the growth of blood-stage *P. vivax*.

Funded by the European Commission and Wellcome Trust; VAC069, VAC071 and VAC079 ClinicalTrials.gov numbers NCT03797989, NCT04009096 and NCT04201431.

## Introduction

*Plasmodium vivax* is the second most common cause of malaria and most geographically widespread, causing an estimated 4.5 million cases in 2020^1^. Control of *P. vivax* is more challenging than *P. falciparum* due to several factors. These include the ability of *P. vivax* to form dormant liver-stage hypnozoites that can reactivate and lead to relapsing blood-stage parasitemia, and earlier production of gametocytes in the blood-stage resulting in more rapid transmission^2^. An effective vaccine would greatly aid elimination efforts worldwide but few *P. vivax* vaccines have reached clinical development.

Candidate vaccines against *P. vivax* have been developed that target different stages of the parasite’s lifecycle^3^. These include blood-stage vaccines that aim to inhibit the invasion of reticulocytes by merozoites, the stage of infection causing clinical disease. The leading blood-stage vaccine target is *P. vivax* Duffy-binding protein (PvDBP), which binds to the Duffy antigen receptor for chemokines (DARC/Fy) on reticulocytes to mediate invasion of the parasite^4^. This interaction is critical as evidenced by the natural resistance of Duffy antigen negative individuals to *P. vivax* malaria^5^. However, the efficacy of blocking this molecular interaction with vaccine-induced antibodies has not been tested previously in clinical trials.

Two vaccines targeting region II of PvDBP (PvDBPII), a 327-amino acid domain that binds to DARC, have previously progressed to Phase I clinical trials. These vaccines comprise a recombinant viral-vectored ChAd63-MVA platform^6^ and a protein/adjuvant formulation (PvDBPII/GLA-SE)^7^. Both vaccines encode the Salvador I (SalI) allele of PvDBPII and were shown to induce binding-inhibitory antibodies (BIA) that block the interaction of recombinant PvDBPII to the DARC receptor *in vitro*^6,7^.

Here we report results from two Phase I/IIa clinical trials in healthy malaria-naïve adults using either the same viral-vectored vaccine or the PvDBPII protein vaccine reformulated in Matrix-M™ adjuvant. Both vaccines were tested for efficacy for the first time by blood-stage CHMI using the heterologous PvW1 clone of *P. vivax*^8^.

## Methods

### Trial design and participants

Two Phase I/IIa vaccine efficacy trials (VAC071, VAC079) and a CHMI trial (VAC069) were conducted in parallel at a single site in the UK (Centre for Clinical Vaccinology and Tropical Medicine, University of Oxford). VAC071 was an open label trial to assess the ChAd63 and MVA viral-vectored vaccines encoding PvDBPII (VV-PvDBPII). The VAC079 trial assessed the protein vaccine PvDBPII in Matrix-M™ adjuvant (PvDBPII/M-M). Efficacy in both trials was determined by impact of the vaccines on PMR following blood-stage CHMI. Unvaccinated infectivity controls, undergoing CHMI in parallel to vaccinees, were enrolled into the VAC069 trial. Eligible volunteers were healthy, Duffy-positive, malaria-naïve adults, aged 18 to 45 years in the vaccine trials and 18 to 50 years in the VAC069 trial. Full details of the inclusion and exclusion criteria are provided in the protocols.

### Trial oversight

The trials were designed and conducted at the University of Oxford and received ethical approval from UK National Health Service Research Ethics Services. The VAC071 and VAC079 vaccine trials were approved by the UK Medicines and Healthcare products Regulatory Agency. All participants provided written informed consent and the trials were conducted according to the principles of the current revision of the Declaration of Helsinki 2008 and ICH guidelines for Good Clinical Practice.

### Vaccines

ChAd63 PvDBPII is a recombinant replication-defective chimpanzee adenovirus serotype 63 and MVA PvDBPII is a modified vaccinia virus Ankara vector, both encoding PvDBPII (SalI allele)^6^. Recombinant PvDBPII protein (SalI allele) was produced in *Escherichia coli* to Good Manufacturing Practices at Syngene International, Bangalore, India^7^. Matrix-M™ is a saponin-based adjuvant provided by Novavax AB, Uppsala, Sweden, which is licensed for use in their COVID-19 vaccine (Nuvaxovid™).

All vaccinations were administered intramuscularly. ChAd63 PvDBPII was administered at a dose of 5×10^10^ viral particles; MVA PvDBPII at 2×10^8^ plaque forming units and PvDBPII protein at 50 μg, mixed with 50 μg Matrix-M™.

### Vaccine safety and immunogenicity

Following each vaccination, local and systemic adverse events (AEs) were self-reported by participants for 7 days. Unsolicited and laboratory AEs were recorded for 28 days after each vaccination. Serious adverse events (SAEs) were recorded throughout the study period. Details on assessment of severity grading and causality of AEs are provided in the protocols. Post-vaccination clinic reviews with hematology, biochemistry and immunology blood tests were conducted at days 1, 3, 7, 14 and 28 after each vaccination. Participants are due to be followed-up to 9 months after their final vaccination. To date, all volunteers have been followed-up to a minimum of 6 months since their final vaccination.

Total anti-PvDBPII IgG serum concentrations were assessed over time by ELISA using standardized methodology^9^. Binding inhibitory antibodies (BIA), which block the interaction of recombinant PvDBPII to DARC *in vitro*, were assessed in serum using an ELISA-based assay^6^. *In vitro* parasite growth inhibition activity (GIA) of 10 mg/mL purified total IgG was measured using a novel transgenic *P. knowlesi* parasite line expressing the PvDBP PvW1 allele (**Fig. S1**), modified from a previous version expressing PvDBP SalI allele^10^. The frequencies of IFN-γ^+^ PvDBPII-specific CD4^+^ and CD8^+^ effector memory T cells were measured using flow cytometry. Details on immunological assays are provided in the Supplementary Appendix.

### Controlled human malaria infection

Vaccinees underwent CHMI 2-4 weeks following their final vaccination and in parallel with unvaccinated infectivity controls in the VAC069 study. Blood-stage CHMI was initiated by intravenous injection of blood infected with the PvW1 clone of *P. vivax*, which originated from Thailand^8^. PvW1 possesses a single copy of the PvDBP gene and its PvDBPII sequence is heterologous to SalI^8^ (**Table S1**). On the day of CHMI, aliquots of 0.5mL cryopreserved PvW1 infected blood were thawed and each participant was challenged with a 1:10 dilution of one aliquot by intravenous injection into the forearm^8^.

From day 6 or 7 post-CHMI, participants were reviewed in clinic once to twice daily for symptoms of malaria and blood parasitemia was measured in real time by qPCR of the 18S ribosomal RNA gene^8^. Volunteers were commenced on antimalarial treatment if they had significant malaria symptoms and parasitemia ≥5,000 genome copies (gc)/mL; or if parasitemia reached ≥10,000 gc/mL irrespective of symptoms. Positive thick film microscopy was also included in the malaria diagnostic criteria in the CHMI trial in 2019 but was removed from later phases (**Fig. 1**). Treatment was with Riamet (60-hour course of artemether/lumefantrine) or Malarone (3-day course of atovaquone/proguanil hydrochloride). Outpatient review continued until completion of antimalarial treatment. Further follow-up visits took place at 2 and 3 months after the day of challenge.

**Figure 1.**
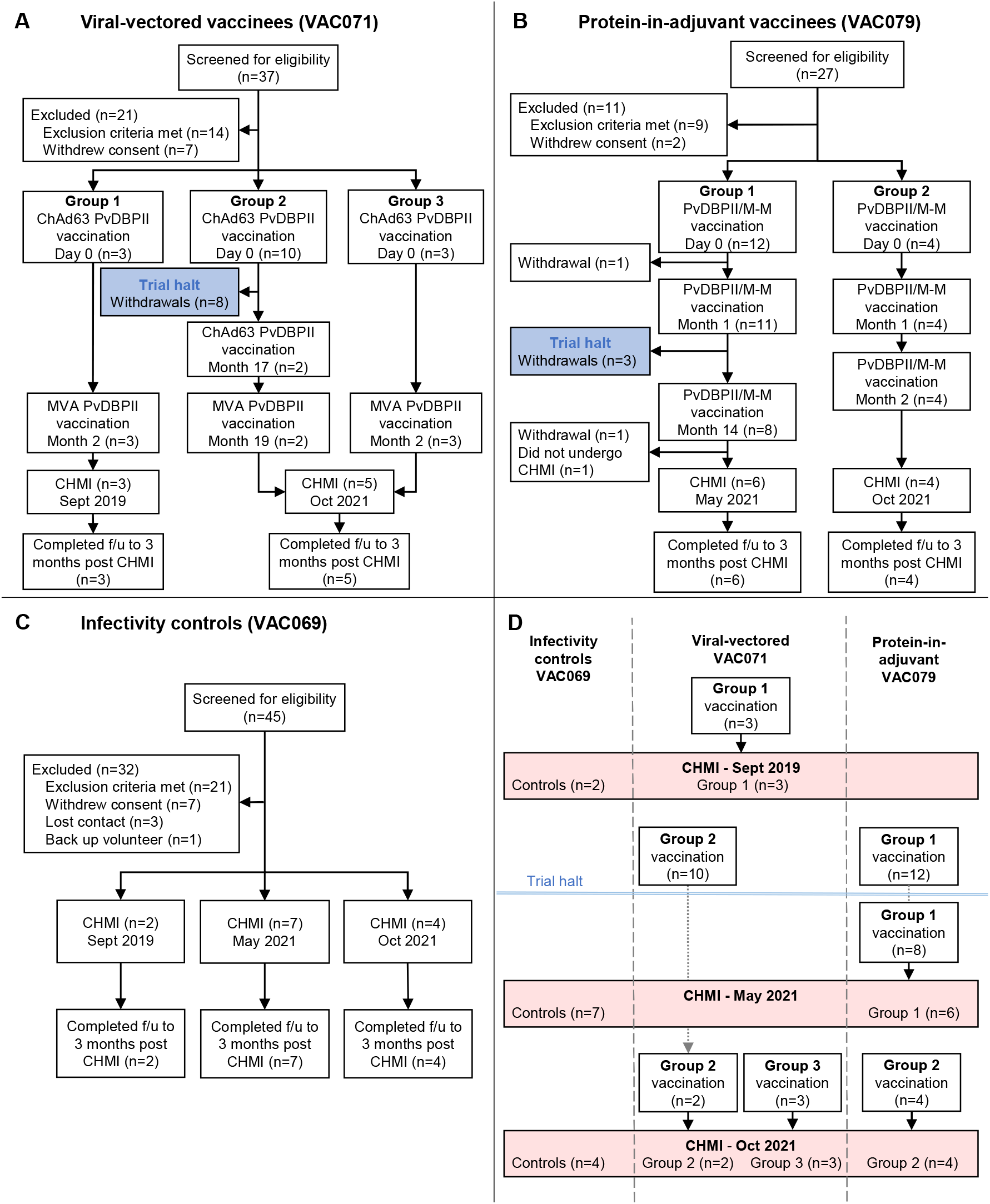
Flow charts of study design and participant recruitment. (**A**): VAC071 Group 1 participants received the viral-vectored vaccines ChAd63 PvDBPII and MVA PvDBPII 8 weeks apart, followed by CHMI 2-4 weeks later. Group 2 received ChAd63 PvDBPII before the trial was temporarily halted. On restart of the trial returning participants in Group 2 received a second dose of ChAd63 PvDBPII at 17 months, followed by MVA PvDBPII 8 weeks later. Group 3 participants received the 8-week viral-vectored vaccine regimen and underwent CHMI along with Group 2 volunteers at 2-4 weeks after the final vaccination. (**B**) VAC079 participants received protein PvDBPII vaccine in Matrix-M™ (M-M) adjuvant. Group 1 volunteers received three doses at 0-1-14 months (delayed third dose due to trial halt). Group 2 volunteers received three doses at 0-1-2 months, with CHMI at 2-4 weeks after the final vaccination. (**C**) VAC069 participants underwent blood-stage CHMI in three separate stages and acted as infectivity controls for vaccinees undergoing CHMI in parallel. (**D**) Summary of the three CHMIs. VAC071 Group 1 vaccinees underwent CHMI in parallel with control participants in September 2019. In January 2020 vaccinations commenced in VAC071 and VAC079, before the trials were halted in March 2020. After restart of the VAC079 trial in 2021, Group 1 participants underwent CHMI in parallel with control participants in May 2021. In October 2021, control participants underwent CHMI in parallel with vaccinees from VAC071 Groups 2 and 3 and VAC079 Group 2.

### Statistical analysis

For the primary efficacy analysis, pairwise comparison of qPCR-derived PMR was made between volunteers who received the same vaccine versus pooled data from all infectivity controls across three CHMIs using Mann-Whitney test. Post-hoc analysis comparing PMR between each vaccination regimen and infectivity controls was performed using Kruskal-Wallis test with Dunn’s multiple comparison post-test. The mean of three replicate qPCR results for each individual at each timepoint was used to model the PMR for each volunteer. PMR was calculated from the slope of a linear model fitted to log_10_ transformed qPCR data^11^. Exploratory analysis of parasite growth was conducted by calculating log_10_ cumulative parasitemia (LCP) for each individual up to the first day on which a volunteer was treated across all CHMIs. Further details of analysis methods are found in the Supplementary Appendix.

Data were analyzed using GraphPad Prism version 8.3.1 for Windows (GraphPad Software Inc) and statistical tests are indicated in the text. Comparisons between groups were performed using Kruskal-Wallis test with Dunn’s multiple comparison post-test. Correlations were assessed using Spearman’s rank correlation. A multiple regression model was used to assess the effect of Duffy blood group serophenotype on PMR, after adjusting for study group. A value of *p*<0.05 was considered significant.

## Results

### Participants

Sixteen volunteers were enrolled into the VAC071 trial testing the viral-vectored vaccines between July 2019 and July 2021 (**Fig. 1A**). Three volunteers in Group 1 received ChAd63 followed by MVA PvDBPII at 0 and 2 months. Ten volunteers in Group 2 received ChAd63 PvDBPII in February 2020, prior to the trial being halted due to the COVID-19 pandemic. After restart of the trial, two of the ten volunteers were re-enrolled and received a second dose of ChAd63 PvDBPII at 17 months, followed by MVA PvDBPII at 19 months. Three volunteers enrolled into Group 3 received one dose of ChAd63 followed by MVA PvDBPII at 0 and 2 months. Vaccinees underwent CHMI 2-4 weeks after their final vaccination.

Sixteen volunteers were enrolled into the VAC079 trial testing PvDBPII/M-M between January 2020 and July 2021 (**Fig. 1B**). Twelve volunteers enrolled into Group 1 in 2020 received two doses of PvDBPII/M-M at 0 and 1 months before the trial was halted due to the COVID-19 pandemic. After restart of the trial in 2021, eight of the twelve volunteers were re-enrolled and received a third vaccination at 14 months and six of these volunteers underwent CHMI 2-4 weeks later. Four volunteers enrolled into Group 2 in July 2021 received three doses of PvDBPII/M-M at 0, 1 and 2 months, followed by CHMI 2-4 weeks later.

Thirteen infectivity control volunteers underwent CHMI in parallel with vaccinees over three phases of the VAC069 study (**Fig. 1C, D**). Demographics of volunteers in each trial are provided in the Supplementary Appendix (**Table S2**).

### Vaccine safety

No safety concerns were identified with the viral-vectored or protein-in-adjuvant vaccines and no SAEs occurred in the VAC071 and VAC079 trials. The viral-vectored vaccines showed similar reactogenicity to that previously reported^6^. Solicited AEs were predominantly mild to moderate in severity, with pain at the injection site and fatigue being most common (**Fig. 2A, B**). Three severe solicited AEs occurred post-vaccination (nausea, feverishness and pyrexia), all of which resolved within 48 hours.

**Figure 2.**
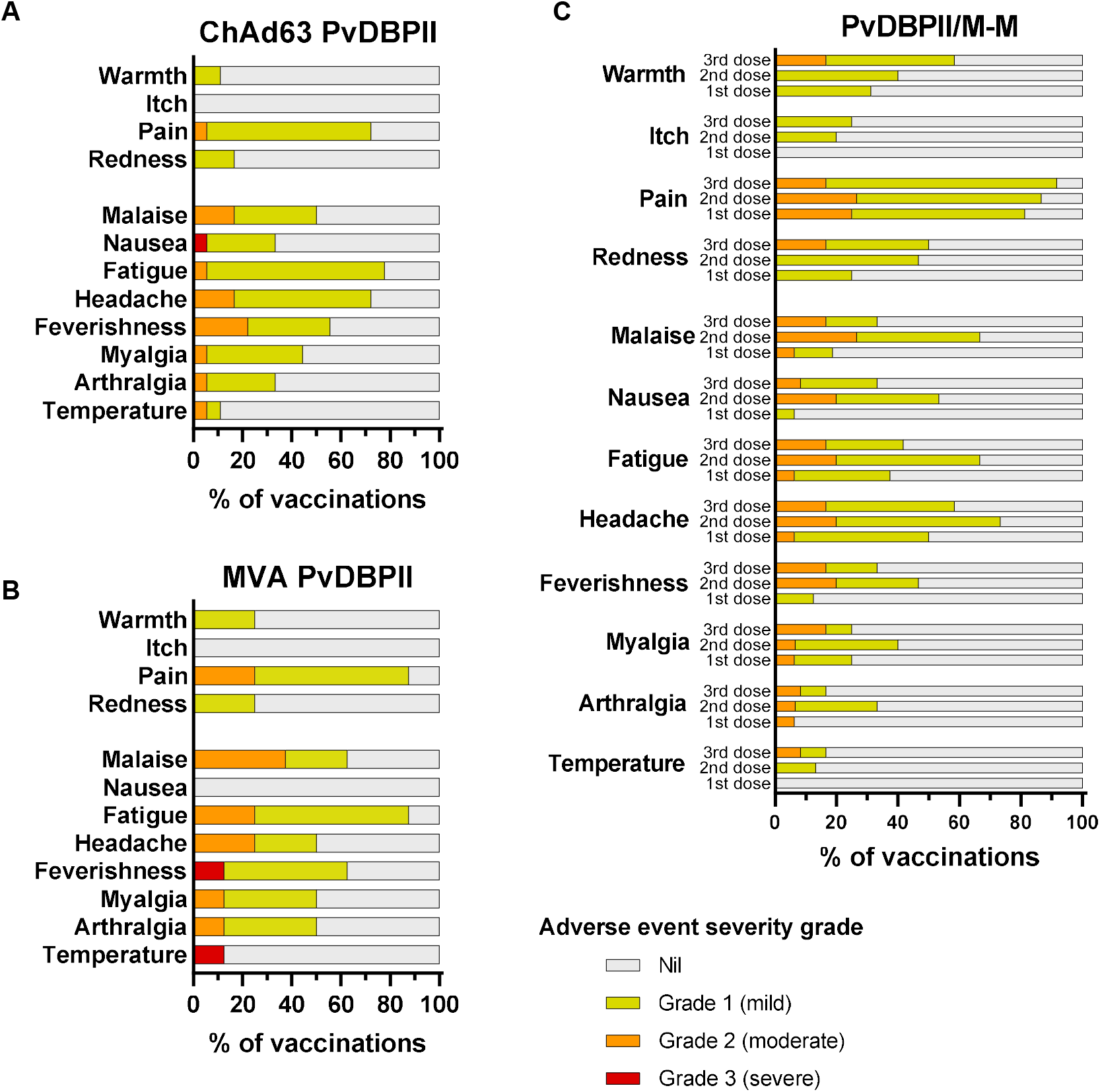
Local and systemic solicited adverse events. Solicited AEs recorded by volunteers within 7 days following each vaccination in participant symptom electronic diaries. The maximal severity reported for each AE is shown as a percentage of the number of vaccinations administered. (**A**) ChAd63 PvDBPII, n=18 vaccinations (16 volunteers received one dose, 2 volunteers received a second dose). (**B**) MVA PvDBPII, n=8 vaccinations (8 volunteers received one dose). (**C**) PvDBPII protein in Matrix-M™ (M-M) adjuvant, AEs reported after first (n=16), second (n=15) and third dose (n=12) are shown.

Solicited AEs following vaccinations with PvDBPII/M-M were all mild to moderate in severity and no severe adverse events occurred (**Fig. 2C**). Injection site pain and headache were the most common solicited AEs.

Transient lymphopenia, with maximal severity of grade 2, occurred commonly following vaccinations with both the viral-vectored and protein-in-adjuvant vaccines (**Table S3**). Unsolicited AEs deemed at least possibly related to either viral-vectored or protein-in-adjuvant vaccinations were of mild to moderate severity and self-limited (**Tables S4, S5**).

### Vaccine immunogenicity

Anti-PvDBPII (SalI) total IgG serum antibody responses peaked around 2 weeks following the final vaccination in all regimens (**Fig. 3A**). PvDBPII/M-M given at 0, 1 and 14 months induced the highest antibody response at this timepoint (geometric mean 198 μg/mL, [range 153-335]), which was significantly higher than the viral-vectored vaccines (29 μg/mL [range 9-85]; *p* <0.001) (**Fig. 3B**). Anti-PvDBPII antibody responses were negative (<1 μg/mL) in all vaccinees prior to their first vaccination, and in controls remained <1 μg/mL throughout.

**Figure 3.**
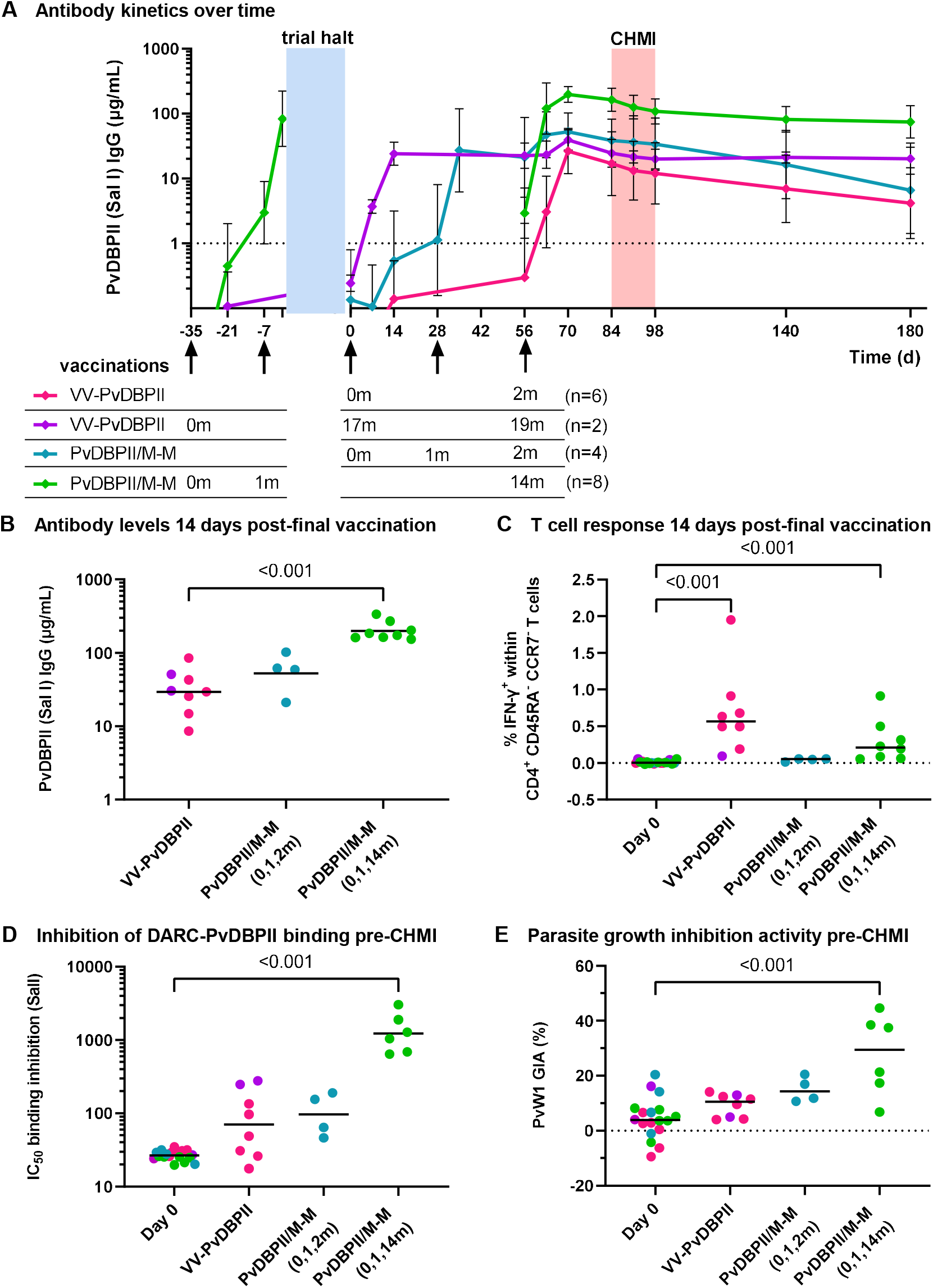
Immunological responses to PvDBPII vaccinations. Anti-PvDBPII Salvador I (Sal I) strain total IgG serum concentrations over time for each vaccination regimen showing geometric mean with standard deviation. Groups are aligned at the time of final vaccination (day 56). Arrows indicate vaccinations with timing of doses in each regimen indicated below in months. VV-PvDBPII = viral-vectored vaccines; PvDBPII/M-M = protein vaccine/Matrix-M™ adjuvant. Blue shading indicates trial halt of ∼1 year, vaccinations occurring prior to the trial halt are shown to the left. Red shading indicates period of controlled human malaria infection (CHMI). IgG concentrations <1 µg/mL, indicated by dashed line, are classified as negative responses but shown for clarity. (**B**) Individual anti-PvDBPII (Sal I) total IgG serum concentrations 14 days post-final vaccination with geometric means for each regimen. (**C**) Percentage of IFN-γ^+^ cells within CD4^+^ CD45RA^-^ CCR7^-^ effector memory T cells 14 days post-final vaccination, following stimulation of peripheral blood mononuclear cells (PBMC) with a pool of PvDBPII peptides. The frequency of IFN-γ^+^ cells in sample-matched unstimulated wells was subtracted to control for non-specific activation. Baseline responses (Day 0) are shown for all volunteers. (**D**) Dilution factor of individual serum, taken pre-CHMI, required to inhibit DARC-PvDBPII (SalI) binding by 50% (IC_50_) with geometric means. Baseline responses (Day 0) are shown for all volunteers. (**E**) Percentage *in vitro* growth inhibition activity (GIA) of 10 mg/mL purified total IgG, taken pre-CHMI, against *P. knowlesi* parasites expressing PvDBP PvW1 allele, with medians. Baseline responses (Day 0) are shown for all volunteers. *p* values as calculated by Kruskal-Wallis test with Dunn’s multiple comparison post-test.

PvDBPII-specific CD4^+^ CD45RA^-^ CCR7^-^ effector memory T cells producing IFN-γ were detectable following final vaccinations with VV-PvDBPII and PvDBPII/M-M administered in a delayed dosing regimen (**Fig. 3C, Table S11**). IFN-γ producing CD8^+^ effector memory T cells were low frequency in the VV-PvDBPII vaccinees and not detectable in the protein vaccine groups (**Figs. S2, S3**).

Serum taken pre-CHMI from vaccinees administered PvDBPII/M-M in the delayed dosing regimen showed ∼10-fold higher levels of BIA (geometric mean of dilution factor to achieve 50% binding inhibition 1224 [range 643-3026]) as compared to the monthly dosing regimen and VV-PvDBPII (**Fig. 3D**). Functional anti-parasitic *in vitro* GIA was generally low pre-CHMI, with the highest levels observed in the PvDBPII/M-M delayed dosing regimen with median GIA of 29% (range 7-45%) (**Fig. 3E**). Serum IgG responses and BIA assayed using the challenge PvW1 sequence of PvDBPII were well correlated and in concordance with responses to the vaccine SalI PvDBPII sequence (**Fig. S4**). BIA also correlated strongly with anti-PvDBPII total IgG serum antibody responses measured by ELISA, whilst GIA versus ELISA indicated the start of a sigmoidal relationship, as previously seen with *P. falciparum* blood-stage vaccines^9^ (**Fig. S5**).

### Vaccine efficacy

Following blood-stage CHMI with the heterologous PvW1 clone of *P. vivax*, all volunteers developed parasitemia and received antimalarial treatment after reaching protocol specified malaria diagnostic criteria (**Fig. 4A, Tables S8-S10**). Volunteers administered the PvDBPII/M-M vaccine, but not VV-PvDBPII, had significantly lower PMR as compared to controls (**Table S6**). Post-hoc analysis showed that this was due to the delayed dosing regimen group of PvDBPII/M-M, who had a significantly lower median PMR of 3.2-fold growth per 48 hours (range 2.3 to 4.3) compared to the unvaccinated controls (median PMR of 6.8-fold growth per 48 hours [range 4.0 to 11.1], *p* <0.001) (**Fig. 4B**). This equated to a 53% reduction in median PMR and was reflected in a 7-day delay in median time to reach malaria diagnosis, from 15.5 days in controls to 22.5 days in vaccinees (**Fig. S6**). Exploratory analysis of log_10_ cumulative parasitemia (LCP) gave concordant results and showed significantly lower LCP in those administered PvDBPII/M-M in the delayed dosing regimen as compared to controls (**Fig. 4C**). PMR and LCP significantly correlated (**Fig. S7**). The other vaccination regimens showed no significant impact on any outcome measure. Parasitemia at the time of malaria diagnosis was consistent across all groups (**Fig. S8**). PMR did not differ by Duffy blood group serophenotype, after adjusting for vaccination group (**Table S7**).

**Figure 4.**
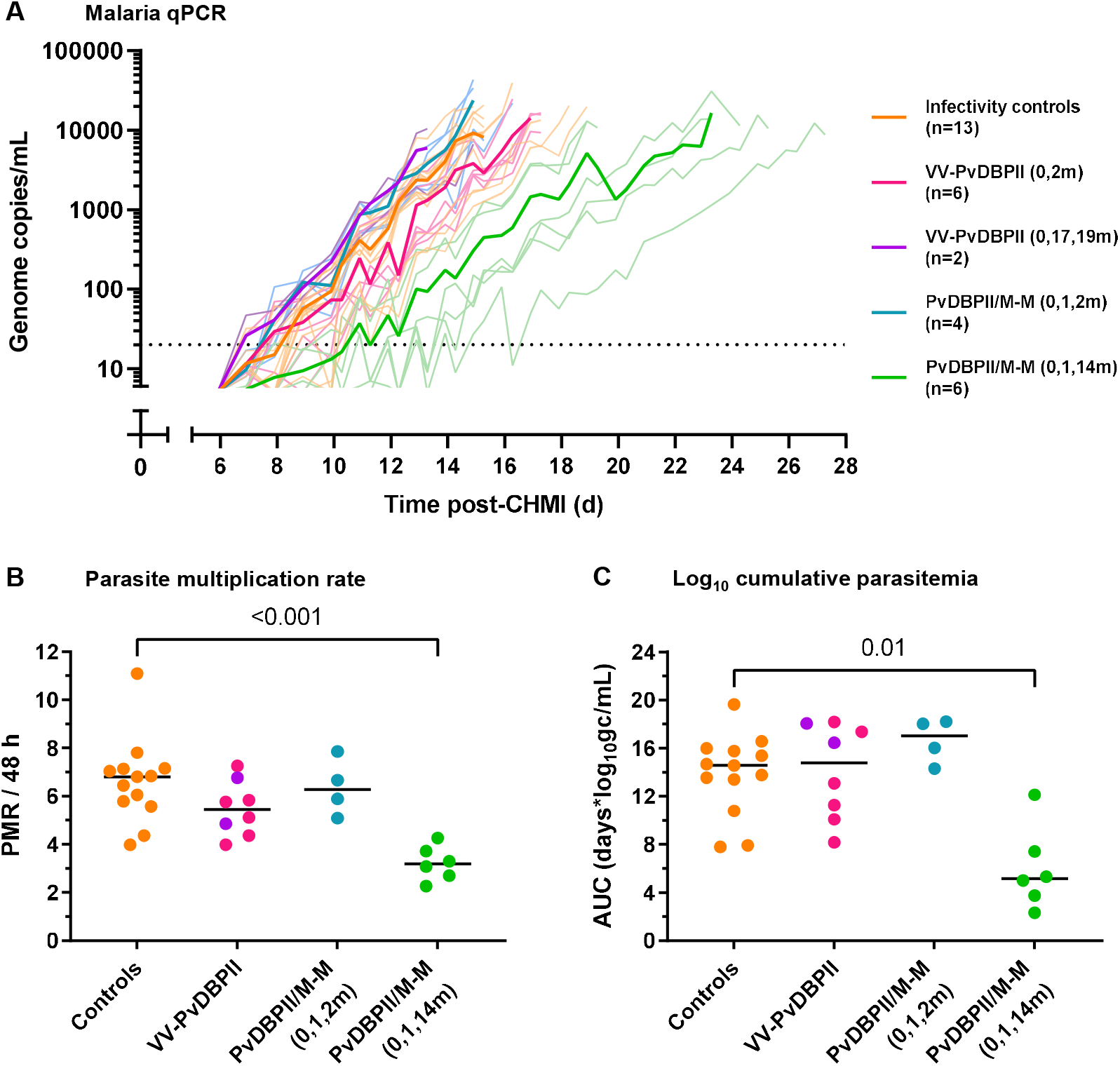
*P. vivax* PvW1 parasitemia after CHMI. (**A**) Individual parasitemia over time measured by qPCR, with group means in bold lines. VV-PvDBPII = viral-vectored vaccines; PvDBPII/M-M = protein vaccine/Matrix-M™ adjuvant. Timings of vaccinations are shown in brackets in months. On the day of CHMI volunteers were administered an intravenous injection of *P. vivax* (PvW1 clone) blood-stage parasites. The dotted line indicates the minimum level of parasitemia to meet positive reporting criteria (20 genome copies [gc]/mL). (**B**) Comparison of parasite multiplication rate (PMR) per 48 hours between vaccinees and controls. Individual PMRs are modelled from the qPCR data over time and are shown with group median. (**C**) Comparison of log_10_ cumulative parasitemia (LCP) during CHMI between vaccinees and controls with group median. LCP calculated from area under the curve (AUC) of log_10_-transformed qPCR over time for each individual, up until day 14 after challenge when the first volunteer reached malaria diagnostic criteria across all CHMIs. *p* values as calculated by Kruskal-Wallis test with Dunn’s multiple comparison post-test.

### Association between immunological readouts and *in vivo* parasite growth inhibition

We assessed the relationship between measurements of vaccine immunogenicity pre-CHMI with *in vivo* growth inhibition (IVGI) observed during CHMI. IVGI was calculated for each vaccinated individual as the percentage reduction in PMR relative to the mean PMR in the unvaccinated controls. The mean IVGI in those administered PvDBPII/M-M in the delayed dosing regimen was 51% (range 36%-66%). We found no association between IVGI and vaccine-induced CD4^+^ T cell IFN-γ responses (**Fig. 5A**). In contrast, correlations were observed between IVGI and all three antibody readouts: anti-PvDBPII (PvW1) total IgG serum antibody ELISA (**Fig. 5B**), BIA using PvW1 sequence PvDBPII protein (**Fig. 5C**) and *in vitro* GIA using purified IgG against *P. knowlesi* parasites expressing the PvDBP PvW1 allele (**Fig. 5D**).

**Figure 5.**
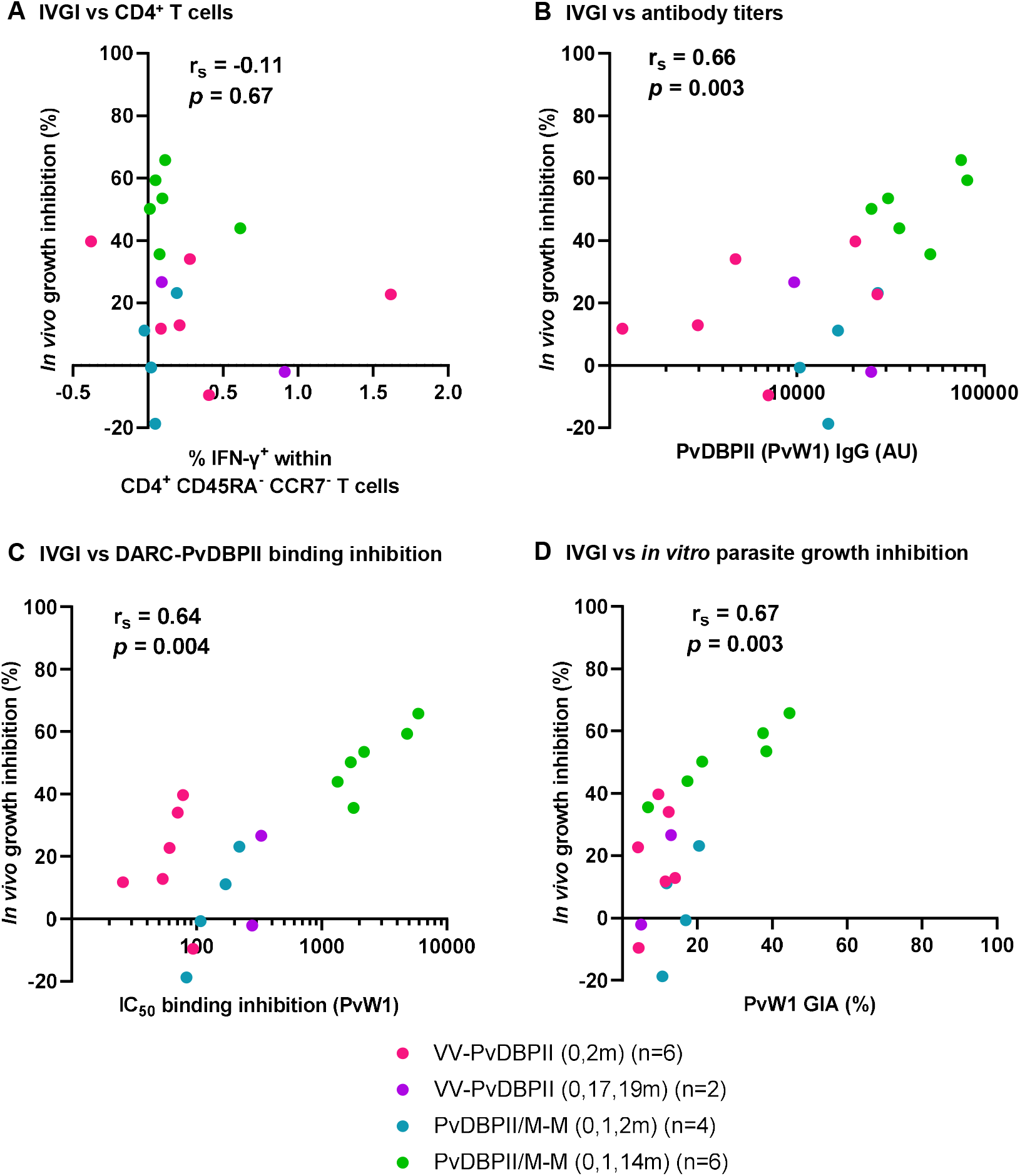
Immune correlates with *in vivo* parasite growth inhibition. Correlation between % *in vivo* parasite growth inhibition (IVGI), calculated as % reduction in PMR in vaccinees relative to the mean PMR in infectivity controls, and pre-CHMI measurements of (**A**) percentage of IFN-γ^+^ cells within CD4^+^ CD45RA^-^ CCR7^-^ effector memory T cells (**B**) anti-PvDBPII (PvW1) total IgG serum titers in arbitrary units (AU); (**C**) dilution factor of individual serum required to inhibit DARC-PvDBPII (PvW1) binding by 50% (IC_50_); and (**D**) % *in vitro* growth inhibition activity (GIA) of 10 mg/mL purified total IgG against *P. knowlesi* parasites expressing the PvDBP PvW1 allele. Spearman’s rank correlation coefficients and *p* values are shown, n=18.

## Discussion

The interaction between PvDBP and its host receptor DARC/Fy is critical for *P. vivax* invasion of reticulocytes, which explains the natural resistance of Duffy-negative individuals to *P. vivax* blood-stage infection^5^. Structural studies have demonstrated that region II within PvDBP binds to DARC^12^ and numerous immuno-epidemiological studies^13,14^ and preclinical vaccine models^15,16^ have supported the hypothesis that vaccine-induced anti-PvDBPII antibodies could prevent blood-stage *P. vivax* parasite growth. Here we present the first clinical vaccine trial confirming this concept.

The Phase I/IIa trials reported here tested two different vaccine platforms to deliver the PvDBPII antigen. Results indicated no safety concerns and both vaccine formulations induced immune responses to PvDBPII. However, following CHMI only the protein-in-adjuvant vaccine PvDBPII/Matrix-M™, given in a delayed 0-1-14 month dosing regimen, inhibited parasite growth. The average reduction of parasite growth by 51% is the largest effect observed to date with any blood-stage malaria subunit vaccine following CHMI and confirms that vaccines targeting PvDBPII can induce significant anti-parasitic immunity.

Previous studies have suggested a role for CD8^+^ T cells in killing of *P. vivax* infected reticulocytes^17^. In this study however, neither vaccine formulation induced a substantial antigen-specific IFN-γ^+^ CD8^+^ T cell response. IFN-γ^+^ CD4^+^ T cell responses to vaccinations were higher, but there was no association between the magnitude of the response with parasite growth during CHMI. We also observed no effect of the Duffy blood group serophenotype on parasite multiplication rate, contrary to reports from field studies^18^, although the number of volunteers in our studies was small. In contrast, our results indicate that the observed anti-parasitic immunity is antibody mediated, as evidenced by the association between IVGI and three *in vitro* readouts of vaccine-induced antibodies: anti-PvDBPII-specific responses (measured by ELISA and functional BIA) and anti-parasitic GIA. These data provide important new benchmarks that link these assay readouts with *in vivo* outcome. The levels of vaccine-induced *in vitro* GIA observed in these trials are modest, in contrast to those recently achieved with optimized blood-stage vaccines for *P. falciparum*^9^. Given that both PvDBPII vaccine candidates tested here were designed over 10 years ago, there is significant potential to rationally improve PvDBP vaccine immunogen design and to identify new blood-stage antigen combinations that elicit higher levels of GIA which would be predicted to confer higher levels of IVGI.

Our results indicate that substantial gains in vaccine-induced antibodies can be achieved via modulation of delivery regimen. The delayed 0-1-14 month dosing regimen with PvDBPII/M-M showed significantly improved immunogenicity, which translated into greater efficacy, as compared to the identical vaccine given in a 0-1-2 monthly regimen. These data add to growing evidence that delayed dosing can improve vaccine-induced antibody responses, as has been seen with a variety of vaccine delivery technologies targeting *P. falciparum* or SARS-CoV-2^9,19-21^ and support further optimization of vaccine regimen to maximize gains in antibody quantity and longevity.

A limitation of our trials is the small number of volunteers in each vaccination group due to withdrawals that occurred during the ∼1 year trial halt secondary to the pandemic, which also necessitated changes to the vaccination regimens partway through the trials. Another limitation is that our studies only used a single clone of *P. vivax* (PvW1) to assess vaccine efficacy. However, the PvW1 clone was recently isolated from a patient in Thailand and thus represents a currently circulating isolate. It also provided a heterologous challenge to the vaccine-induced responses raised against the SalI allele of PvDBPII. The efficacy results in these trials indicate that human immunization with this immunogen can raise antibodies that recognize conserved epitopes within diverse PvDBPII variants. It will nonetheless be important for future studies to test the efficacy of PvDBPII-based vaccines against other heterologous *P. vivax* strains from different geographic locations, strains with PvDBP gene copy number variation^22^, and parasites that infect Duffy-negative individuals^23^.

Overall this study represents a milestone for the *P. vivax* blood-stage malaria vaccine field by confirming that vaccine-induced anti-PvDBPII immune responses can impact *P. vivax* growth in malaria-naïve individuals *in vivo*. Next steps will include CHMI or field efficacy trials of PvDBPII/M-M in malaria-endemic populations to explore whether this vaccine can enhance pre-existing anti-malarial antibody responses. In parallel, avenues to improve vaccine efficacy should be explored, including combining this vaccine with those targeting other lifecycle stages^24^ and optimizing new blood-stage vaccines. Our data provide the framework, with defined benchmark levels of anti-PvDBPII antibodies and GIA versus IVGI, to guide rational design and delivery of next-generation blood-stage vaccines to protect against *P. vivax* malaria.

## Data Availability

All data produced in the present study are available upon reasonable request to the authors

## Acknowledgments

The authors are grateful for the assistance of Aabidah Ali, Megan Baker, Duncan Bellamy, Nicholas Byard, Federica Cappuccini, Hannah Davies, Francesca Donnellan, Amy Flaxman, Julie Furze, Michelle Fuskova, Susanne Hodgson, Daniel Jenkins, Kimberly Johnson, Kathryn Jones, Reshma Kailath, Colin Larkworthy, Fernando Ramos Lopez, Meera Madhavan, Rebecca Makinson, Daniel Marshall-Searson, Celia Mitton, Abigail Platt, Jack Quaddy, Raquel Lopez Ramon, Indra Rudiansyah, Hannah Scott, Merin Thomas, Cheryl Turner, Nicola Turner, Marta Ulaszewska, Chris Williams, Rhea Zambellas (Jenner Institute, University of Oxford); Robert Smith, Eleanor Berrie, Helena Parracho and Richard Tarrant (Clinical Biomanufacturing Facility, University of Oxford); Sally Pelling-Deeves for arranging contracts (University of Oxford); Julie Staves and the Haematology Laboratory (Oxford University Hospitals NHS Foundation Trust); Xin Xue Liu for statistical advice (Oxford Vaccine Group, University of Oxford); Karin Lövgren Bengtsson (Novavax); Nongnuj Maneechai, Tianrat Piteekan, Nattawan Rachaphaew, Wanlapa Roobsoong, Jetsumon Sattabongkot, Nick Day (Mahidol Vivax Research Unit, Thailand); Wai-Hong Tham, Meta Roestenberg, and Susan Barnett for providing scientific advice as part of the MultiViVax Scientific Advisory Board; past members of the Malaria Group, ICGEB including Sanjay Singh, Kailash Pandey, Shams Yazdani, Ahmed Rushdi Shakri, Dhiraj Hans and Rukmini Bharadwaj and past members of MVDP, including Kavita Singh, Gaurav Pandey, Shantanu Mehta and Rajender Jena, for playing key roles in development of PvDBPII; and all the study volunteers.

## Author Contributions

- Designed the study: MMH, YT, TAR, SES, CEC, AMM, SJD.
- Collected the data: MMH, JRB, YT, TAR, AD, FJM, CMN, AML, LDWK, NJE, NMG, LK, IDP, BK, CG, DJML, JS, MG-B, KM, SES, AMM.
- Analyzed the data: MMH, JRB, CMN, NJE, KM, SES, CEC, AMM, SJD.
- Contributed reagents / materials / analysis tools: CH, JMR, VSC, PM, CAL, FM, RWM, KM, CEC.
- Project Management: PM, SB, IJT, AML, JSC, FLN.
- Wrote the paper: MMH, JRB, AMM, SJD.

