## supplemental appendix for "Impact of a blood-stage vaccine on *Plasmodium vivax* malaria"

### Supplementary Appendix

#### Supplementary Methods

##### Trial approvals

The studies received ethical approval from UK National Health Service Research Ethics Services, (VAC069: Hampshire A Research Ethics Committee, Ref 18/SC/0577; VAC071: Oxford A Research Ethics Committee, Ref 19/SC/0193; VAC079: Oxford A Research Ethics Committee, Ref 19/SC/0330). The vaccine trials were approved by the UK Medicines and Healthcare products Regulatory Agency (VAC071: EudraCT 2019-000643-27; VAC079: EudraCT 2019-002872-14).

##### Peripheral Blood Mononuclear Cell (PBMC), plasma and serum preparation

Blood samples were collected into lithium heparin-treated vacutainer blood collection systems. PBMC were frozen in foetal calf serum containing 10% dimethyl sulfoxide and stored in liquid nitrogen. Plasma samples were stored at -80 °C. For serum preparation, blood samples were collected into untreated vacutainers, incubated at room temperature and then the clotted blood was centrifuged for 5 min (750 *xg*). Serum was stored at -80 °C.

##### Anti-PvDBPII standardized ELISA

ELISAs to quantify circulating PvDBPII-specific total IgG responses were performed using standardized methodology, similar to that previously described[1]. Nunc MaxiSorp ELISA plates (Thermo Fisher) were coated overnight (≥16 h) at 4 °C with 50 µL per well of 2 µg/mL PvDBPII (SalI or PvW1 allele) protein[1]. Plates were washed 6x with 0.05 % PBS/Tween (PBS/T) and tapped dry. Plates were blocked for 1 h with 100 µL per well of Starting Block™ T20 (Thermo Fisher) at 20 °C. Test samples were diluted in blocking buffer (minimum dilution of 1:100), and 50 µL per well was added to the plate in triplicate. Reference serum (made from a pool of high-titer vaccinated donor serum) was diluted in blocking buffer in a three-fold dilution series to form a ten-point standard curve. Three independent dilutions of the reference serum were made to serve as internal controls. The standard curve and internal controls were added to the plate at 50 µL per well in duplicate. Plates were incubated for 2 h at 20 °C and then washed 6x with PBS/T and tapped dry. Goat anti-human IgG–alkaline phosphatase secondary antibody (Merck) was diluted 1:1000 in blocking buffer and 50 μL per well was added. Plates were incubated for 1 h at 20 °C. Plates were washed 6x with PBS/T and tapped dry. 100 μL per well of PNPP alkaline phosphatase substrate (Thermo Fisher) was added, and plates were incubated for approximately 15 min at 20 °C. Optical density at 405 nm ($OD_{405}$) was measured using an ELx808 absorbance reader (BioTek) until the internal control reached an $OD_{405}$ of 1.0. The reciprocal of the internal control dilution giving an $OD_{405}$ of 1.0 was used to assign an arbitrary unit (AU) value of the standard. Gen5 ELISA software v3.04 (BioTek) was used to convert the $OD_{405}$ of test samples into AU by interpolating from the linear range of the standard curve fitted to a four-parameter logistic model. Any test samples with an $OD_{405}$ below the linear range of the standard curve at the minimum dilution tested were assigned a minimum AU value of 5.0. These responses in AU are reported in μg/mL for the PvDBPII SalI allele following generation of a conversion factor by calibration-free concentration analysis (CFCA) as reported previously[1].

**ELISA-based Binding Inhibition Assay**

Samples were analysed for binding inhibitory antibodies (BIA) at the Institut Pasteur, Paris, using previously reported methodology[2]. Recombinant DARC-Fc (1 μg/mL) was coated on to a 96-well plate overnight at 4 °C in carbonate-bicarbonate buffer. Next day, the plate was blocked for 2 h at 37 °C using 2 % non-fat milk. Recombinant PvDBPII (SalI or PvW1 sequence) in a range of 0.8–25 ng/mL was used to generate a PvDBPII standard curve using a four-parameter logistic model. Serum samples were analysed at dilutions of 1:10 to 1:2430. Each serum dilution was incubated with 25 ng/mL PvDBPII protein at 37 °C for 30 min. The reaction mixture was then added to DARC-Fc coated wells of an ELISA plate and incubated at 37 °C for 1 h. PvDBPII protein bound to recombinant DARC was probed with anti-PvDBPII polyclonal rabbit sera at 37 °C for 1 h and detected with anti-rabbit IgG HRP-conjugated secondary antibody at 37 °C for 1 h. The assay was developed using the two-component chromogenic substrate for peroxidase detection TMB (3,3′,5,5′-tetramethylbenzidine, Life Sciences) for 5 min and the reaction was stopped with phosphoric acid 1M

($H_3PO_4$). Absorbance was immediately measured at a wavelength of 450 nm. The amount of bound PvDBPII was estimated by converting OD values to protein concentrations using the PvDPBII standard curve. The interpolated protein concentration values were used to calculate percent binding for each serum sample dilution. The % binding inhibition for each serum dilution was calculated as follows: % Binding Inhibition = 100 - % Binding. The plot of % Binding Inhibition versus serum dilution was used to find the serum dilution at which 50% binding inhibition ($IC_{50}$) was achieved. Each assay was performed in duplicate and results from three independent replicates were used to determine average $IC_{50}$.

***P. knowlesi* parasites and growth inhibition activity (GIA) assay**

It is not possible to culture blood-stage *P. vivax* long-term *in vitro* and therefore *P. vivax* parasites cannot be used at scale in GIA assays. Instead a transgenic *P. knowlesi* (a closely related simian malaria species) parasite line was generated, which is adapted to long-term *in vitro* culture in human red blood cells. We previously generated a transgenic *P. knowlesi* parasite line (*P. knowlesi* $PvDBP^{OR}\Delta\beta\Delta\gamma$), in which the PvDBP SalI allele transgene replaced the native PkDBPα gene, and the PkDBPβ and PkDBPγ genes were also knocked out[3]. Here, we further modify this line to create a novel transgenic *P. knowlesi* line expressing the PvDBP PvW1 allele (**Fig. S1**). A 20 bp guide sequence (CGA CAT CCT GAA GCA GGA AC) targeting the recodonized PvDBP (SalI) was identified and cloned into the PkCas9/sgRNA vector ($pCas9/sg\_PvDBPRII(SalI)^{OR}$) as previously described[3-5]. A donor plasmid, pDonor_PvW1, was created by cloning a synthetic recodonized PvDBP PvW1 allele (GeneArt, Thermofisher) into the plasmid $pDonor\_PkDBP\alpha^{OR}$, via SpeI and NotI restriction sites. This created the final vector containing PvDBP PvW1 gene sequence flanked by 5' and 3' homology regions targeting the PkDBPα locus. The donor and guide were transfected into *P. knowlesi* $PvDBP^{OR}\Delta\beta\Delta\gamma$ using previously described methods[3]. The resultant transfectants were cloned via limiting dilution and genotyped by PCR as described previously using diagnostic oligos for WT $PvDBPRII^{OR}$ locus (ol186 fwd-CAC GAT TTG TGT ACT TAT AGA ATC AAT TTT TCC TT and ol189 rev-CGT TCT GGC CGT CGC CTG T) and for successful integration of PvDBP PvW1 allele (ol186 fwd and ol1799 rev-TCC CGT TCT TCC CAT CTC CGG T).

Samples were analysed by the GIA Assay Reference Center, Laboratory of Malaria and Vector Research, National Institute of Allergy and Infectious Diseases, National Institutes of Health, US. The GIA assay methodology has been published elsewhere[6]. In brief, 10 mg/mL purified total IgG (Protein G purified from serum) were mixed with ~1.5 % of trophozoite-rich parasites in a final volume of 40 μL in 96-well plates. After ~27 h of incubation, the relative parasitemia in each well was determined by parasite lactate dehydrogenase (pLDH) activity. Test IgG samples which showed >10% GIA in the first assay were tested in two more independent assays, and median % GIA values from the three assays are reported.

**Flow cytometry T cell assay**

Flow cytometry was performed using frozen aliquots of PBMC from donors on day 0 (pre-vaccination) and 14 days and 28 days post-final vaccination with either VV-DBPII or PvDBPII/M-M. Cryopreserved PBMC were thawed and rested before an 18 h stimulation with medium alone, 2.5 μg/peptide/mL of a PvDBPII 20mer peptide pool (Mimotopes) (**Table S11**), or 1 μg/mL Staphylococcal enterotoxin B (SEB; S-4881, Sigma; positive control). Anti-CD28 (1 μg/mL; 16-0289-85, eBioscience, clone: CD28.2), anti-CD49d (1 μg/mL; 16-0499-85, eBioscience, clone: 9F10) and anti-CD107a-PE-Cy5 (15-1079-42, eBioscience, clone: eBioH4A3) were included in the cell culture medium. Brefeldin A (00-4506-51, eBioscience) and monensin (00-4505-51, eBioscience) were added after 2 h. Following incubation, PBMC were stained and fixed with Cytofix/Cytoperm (554714, BD Biosciences). The following anti-human antibodies / dyes were used: anti-CD14-eFluor™ 450 (48-0149-42, clone: 61D3), anti-CD19-eFluor™ 450 (48-0199-42, clone: HIB19), anti-CD8a-APC-eF780 (47-0088-42, clone: RPA-T8), anti-IFN-γ-FITC (11-7319-82, clone: 4S.B3), anti-TNFα-PE-Cyanine7 (25-7349-82, clone: MAb11), anti-CD3-Alexa Fluor® 700 (56-0038-82, clone: UCHT1) – all eBioscience; anti-CD4-PerCP Cy5.5 (300530, clone: RPA-T4), anti-IL-2-BV650 (500334, clone: MQ1-17H12), anti-IL5-PE (500904, clone: JES-39D10), anti-IL13-APC (501907, clone: JES10-5A2), anti-CD45RA-BV605 (304134, clone: HI100), anti-CCR7-BV711 (353228, clone: G043H7) – all Biolegend; and Live/Dead Aqua (L34966, Invitrogen). Samples were acquired on a Fortessa flow cytometer using BD FACSDiva (both BD Biosciences) and data were analyzed in FlowJo (v10.8, Treestar).

**Blood-stage inoculum preparation and CHMI**

The PvW1 blood-stage inoculum was thawed and prepared under strict aseptic conditions as previously described[7]. The required number of vials of the cryopreserved stabilate (each containing approximately 0.5 mL of red blood cells in 1 mL of Glycerolyte 57) were thawed in parallel in an area using solutions licensed for clinical use and single-use disposable consumables. A class II microbiological safety cabinet (MSC) was used to prepare the inoculum, which was fumigated with hydrogen peroxide and decontamination validated prior to use. To prepare the inoculum, 0.2 volume 12 % saline was added dropwise to the contents (~1.5 mL) of each vial of thawed infected blood. Each sample was left for 5 min, before an additional 10 volumes of 1.6 % saline was added dropwise prior to centrifugation for 4 min at 830 x*g*. Each supernatant was removed and 10 mL of 0.9 % saline was added dropwise. The cell pellets were pooled and washed twice in 0.9 % saline before a final resuspension into one 10mL sample in 0.9 % saline. This 10 mL suspension was then divided into aliquots, equivalent to one tenth of one original cryovial. Each aliquot was made up to a total volume of 5 mL in 0.9 % saline in a sterile syringe for injection and transported to the clinic.

The reconstituted blood-stage inoculum (5 mL per syringe) was injected intravenously via an indwelling cannula, preceded and followed by a saline flush. The inoculum was administered to all volunteers within a maximum of 3 h 7 min from thawing of the inoculum. Volunteers were observed for 1 h following injection of the inoculum before discharge from the clinical facility. Following each CHMI, a leftover sample of the inoculum was cultured and shown to be negative for bacterial contamination.

**Malaria parasite quantification by qPCR**

Quantitative PCR (qPCR) was used to measure *P. vivax* parasitemia in volunteers' blood in real-time as previously described[7] using an assay that targets the 18S ribosomal RNA (rRNA) gene. DNA was extracted from 0.4 mL whole EDTA blood using a QIAsymphony SP robot, utilizing the Qiagen DSP Blood Midi Kit and the pre-loaded Blood 400 v6 extraction protocol, with a 100 μL elution in ATE buffer selected. Additionally, aliquots of baseline samples taken within 2 days pre-CHMI were spiked with a known concentration of positive control DNA to check there was no presence of PCR inhibitors in volunteers' blood prior to CHMI.

Following DNA extraction, a standard Taqman absolute quantitation was used against a standard curve to amplify a 183 bp PCR product from the multi-copy, highly conserved 18S ribosomal RNA genes of *Plasmodium spp.* qPCR used the following adapted oligonucleotide primers and probe[8]: 18s forward primer 5'-AGG AAG TTT AAG GCA ACA ACA GGT-3', 18s reverse primer 5'-GCA ATA ATC TAT CCC CAT CAC GA-3' and shortened FAM labelled probe sequence 5'-TGA ACT AGG CTG CAC GCG-3', was run on an ABI StepOne Plus machine with v2.3 software. Default Universal qPCR (target FAM-NFQ-MGB) and QC settings were used apart from the use of 40 cycles and 25 μL reaction volume.

This qPCR detects DNA from pan-*Plasmodium* species, but unlike the synchronous growth of *P. falciparum*, circulating *P. vivax* infected RBC may contain up to 10-15 individual genomes (in blood-stage late trophozoites and schizonts) and can also include the presence of gametocytes. The qPCR score is therefore reported in genome copies/mL (gc/mL) as opposed to a quantity of parasites.

The standard curve was generated from dilution of a linearized plasmid encoding part of the *Plasmodium spp.* 18S ribosomal RNA gene and calibrated using known *P. falciparum* (Pf) spiked blood samples initially and then reference DNA extracted from whole blood from *P. vivax*-infected patient samples in Thailand where parasites had been quantified by microscopy (kindly provided by Mahidol University). Based upon earlier results obtained using dilution series of microscopically-counted cultured Pf parasites, a Pf-specific 18S rRNA Taqman qPCR showed a lower limit of quantification (LLQ, defined as %CV <20%) of around 20 Pf parasites (p)/mL blood[9]. Counted parasite dilution series results also suggested that the lower limit of probable detection (LLD, i.e. a probability of >50% of ≥1 positive result among three replicate qPCR reactions) is in the region of 5 p/mL, while samples at 1 p/mL are consistently negative (24/24 qPCR reactions). Positive results in this assay (even at very low level) are thus essentially 100 % specific for genuine parasitemia, with positive results beneath the LLQ likely to signify parasitemia in the range 2-20 p/mL. Similar sensitivity in terms of genome copy detection was observed when using the pan-*Plasmodium* qPCR described above and the diluted *P. vivax*-infected patient blood test samples from Thailand. As noted, these samples had microscopically mixed life stages with varying copies of the 18S rRNA gene and thus the assay readout is reported in terms of gc/mL. Based on this and the above experiments, 20 gc/mL was set as the minimum level to meet positive reporting criteria, but all raw data are shown in the Results.

For quality control purposes, qPCR samples were re-tested if;

- Replicates included a mixture of positive and negative (in terms of amplification) results with one or more positive results > 100 gc/mL;
- The % CV of any results were high outliers.

All 'passed' data following the quality control steps above, including any 0 values, were used to generate the final mean qPCR result for each time-point.

**Thick blood film microscopy**

Collection of blood, preparation of thick films and slide reading were performed according to Jenner Institute Standard Operating Procedure (SOP) ML009. Slides were prepared using Field's stain A and then Field's stain B. 200 fields at high power (1000x) were read. Visualization of 2 or more parasites in 200 high power fields constituted a positive result. For internal quality control, all slides were read separately by two experienced Thai microscopists, with a third read if results were discordant.

**Modelling of parasite multiplication rate**

A qPCR-derived parasite multiplication rate (PMR) was modelled based on previously described methodology with modifications[7,9]. The arithmetic mean of three replicate qPCR results obtained for each individual at each time-point was used for model-fitting. Negative replicates and any qPCR data points < 20 gc/mL, based upon the mean of the three replicates, were removed prior to model-fitting. Data from timepoints in CHMIs conducted in September 2019 and May 2021, which would not have been available if using the visit schedule for the final CHMI in October 2021 (VAC069D /VAC071B /VAC079B), were also removed prior to model-fitting. The time interval between the morning and evening bleeds used for qPCR monitoring was set as 0.37 days. PMR per 48 h was then calculated using a linear model fitted to $\log_{10}$-transformed qPCR data.

**Analysis of $\log_{10}$ cumulative parasitemia during CHMI**

The arithmetic mean of three replicate qPCR results obtained for each individual at each time-point up until day C+14, when the first volunteer reached malaria diagnostic criteria across all CHMIs, was used for analysis. As per PMR modelling, negative replicates and any qPCR data points < 20 gc/mL, based upon the mean of the three replicates, were removed prior to analysis, as well as removal of data from timepoints in CHMIs conducted in September 2019 and May 2021, which would not have been available if using the visit schedule for the final CHMI in October 2021. $\text{Log}_{10}$ cumulative parasitemia was then calculated from area under the curve analysis of $\log_{10}$-transformed qPCR data, where a peak was defined as any positive value above baseline.

#### Supplementary Figures

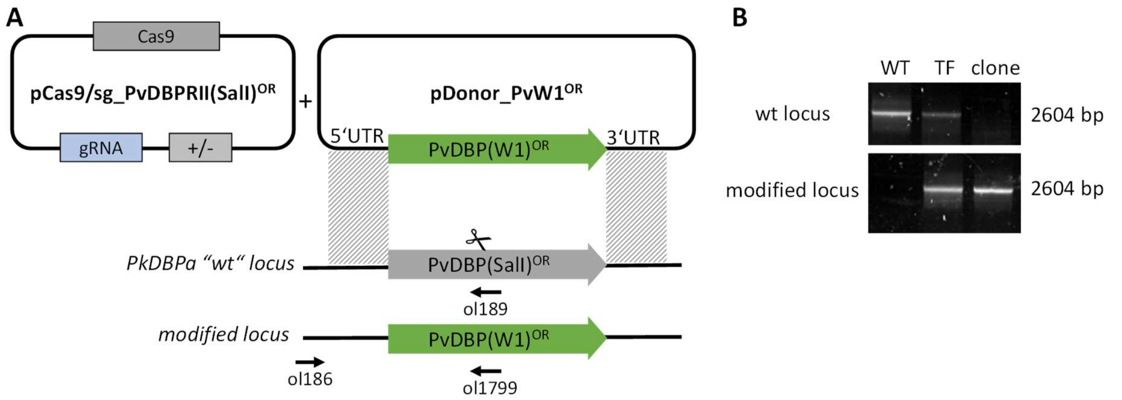

**Figure S1. Design and genotypic analysis of *P. knowlesi* PvDBP PvW1 orthologue replacement line.**

(**A**) Schematic detailing the approach to replace the coding sequence of PvDBP SalI allele with that of the PvDBP PvW1 allele within the previously generated transgenic *P. knowlesi* PvDBP$^{OR}\Delta\beta\Delta\gamma$ ("wild type" WT) strain. The pCas9/sg_PvDBPRII(SalI)$^{OR}$ plasmid was transfected alongside the pDonor_PvW1$^{OR}$, to create a double strand break within the PvDBP SalI coding sequence and replace this with the PvDBP PvW1 allele by homologous recombination. Arrows indicate positions of diagnostic primers used for genotypic analysis of transfectants. (**B**) Parasites were analysed by diagnostic PCR. Primer pairs were used to specifically detect wt locus (ol186 + ol189) and the modified locus (ol186 +ol1799) within i) the parental *P. knowlesi* PvDBP$^{OR}\Delta\beta\Delta\gamma$ (WT) strain; ii) bulk culture of transfectants (TF); and iii) a clonal transfectant (clone). The clonal *P. knowlesi* PvDBP$^{OR}$ PvW1 transgenic parasite line was shown to only contain parasites with the PvW1 allele modified locus.

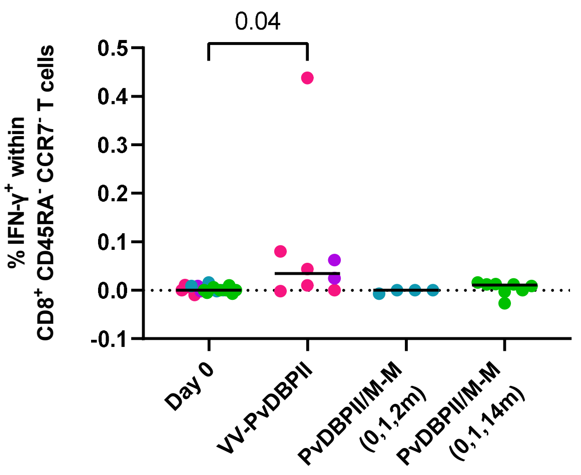

**Figure S2. PvDBPII-specific CD8$^+$ T cell responses 14 days post-final vaccination.**

Percentage of IFN-γ$^+$ cells within CD8$^+$ CD45RA$^-$ CCR7$^-$ effector memory T cells 14 days post-final vaccination following PBMC stimulation with a pool of PvDBPII peptides. The frequency of IFN-γ$^+$ cells in sample-matched unstimulated wells was subtracted to control for non-specific activation. Baseline responses (Day 0) are shown for all volunteers. *p* value as calculated by Kruskal-Wallis test with Dunn's multiple comparison post-test.

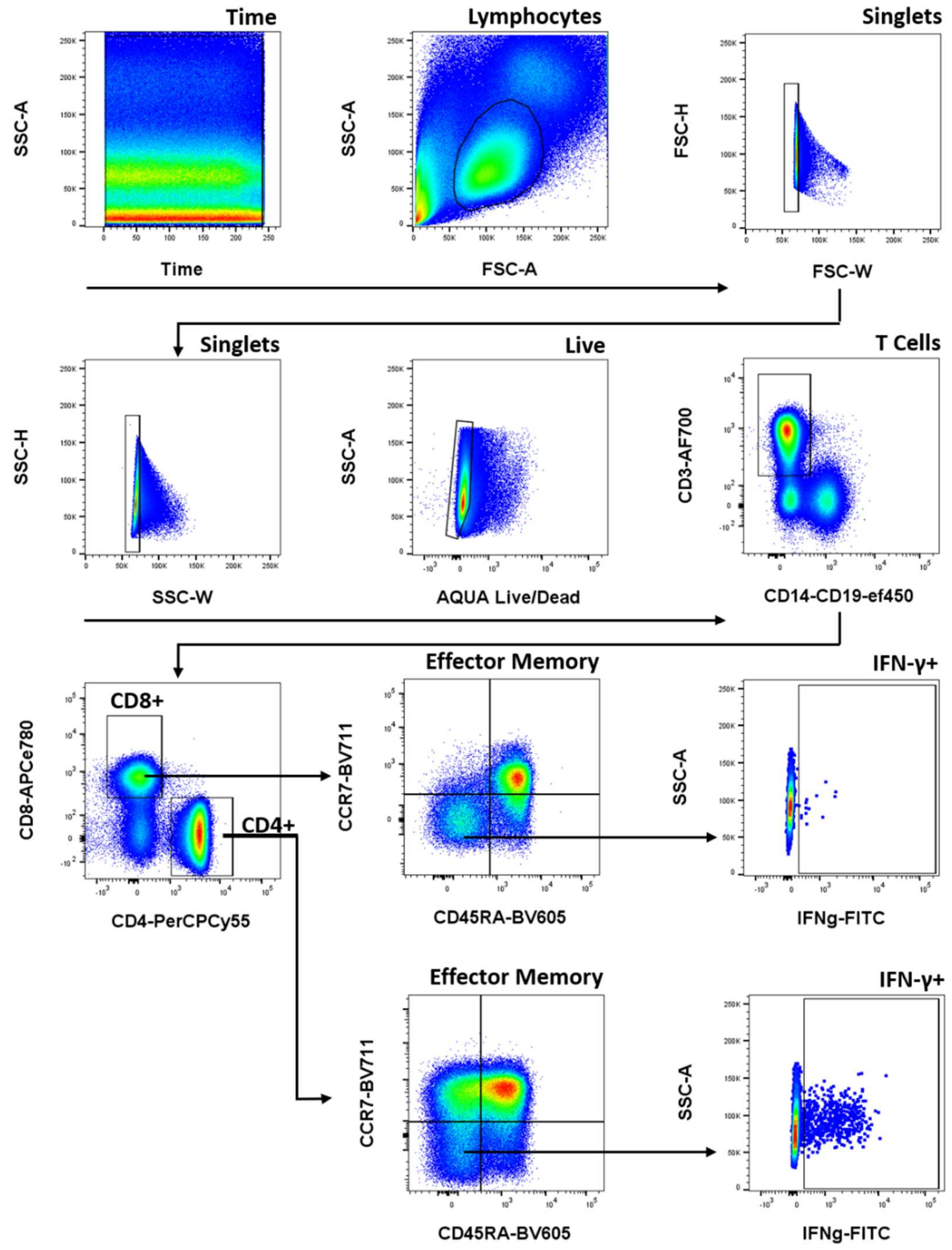

**Figure S3. Flow cytometry gating strategy.**

Gating strategy for definition of live singlet $CD4^+$ and $CD8^+$ effector memory T cells, and for gating of IFN-$\gamma^+$ cells within the live singlet $CD4^+$ and $CD8^+$ effector memory T cell populations.

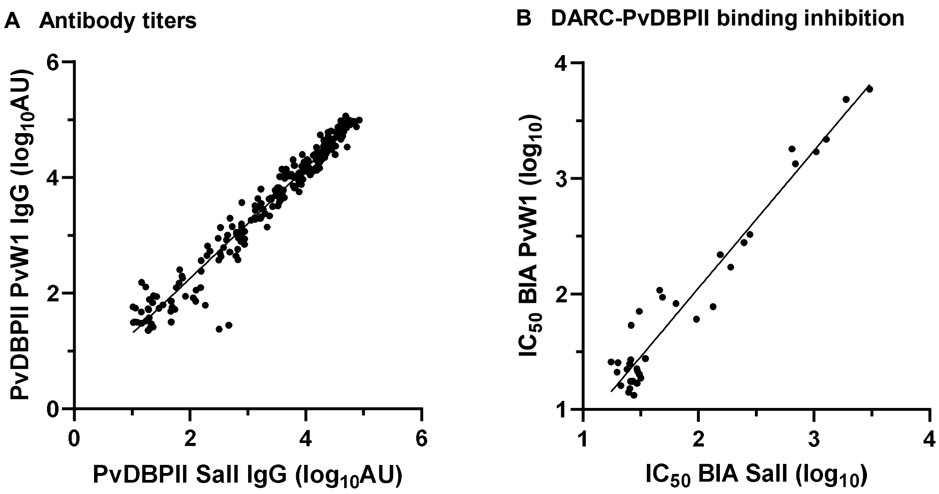

| | Antibody titers | Binding inhibition |
|---|---|---|
| **Slope** | 0.95 | 1.19 |
| **95% CI** | 0.93 to 0.98 | 1.09 to 1.29 |
| **$R^2$** | 0.95 | 0.94 |
| **n** | 230 | 36 |

**Figure S4. Analysis of anti-PvDBPII SalI versus PvW1 ELISA and BIA responses.**

(**A**) ELISA data from all volunteers and timepoints at which anti-PvDBPII total IgG responses were assessed in serum. Responses to both variants of PvDBPII are reported in $log_{10}$ arbitrary units (AU). Linear regression line is shown. (**B**) Data from all volunteers and timepoints at which BIA in serum were assessed via inhibition of recombinant DARC-PvDBPII binding. $Log_{10}$ dilution factor of individual serum required to inhibit binding of DARC to both variants of PvDBPII by 50% ($IC_{50}$) are reported. Linear regression line is shown.

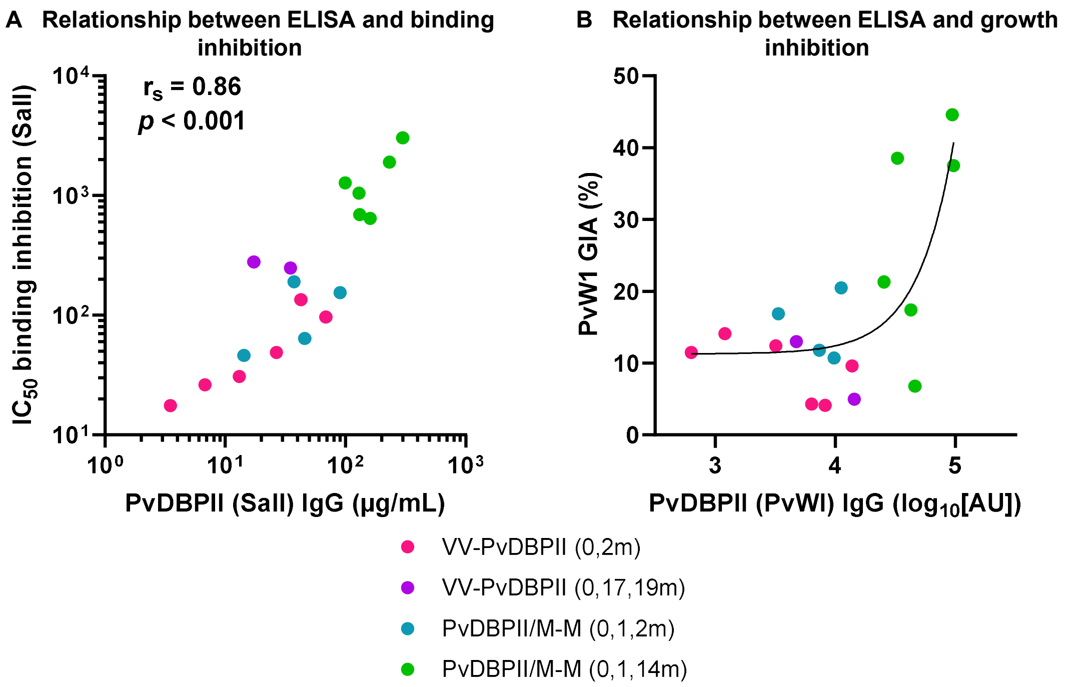

**Figure S5. Relationships between measures of anti-PvDBPII antibody responses pre-CHMI.** (**A**) Correlation analysis between anti-PvDBPII (SalI) total IgG serum responses measured pre-CHMI by ELISA versus BIA measured at the same timepoint by dilution factor of individual serum required to inhibit DARC-PvDBPII (SalI) binding by 50% ($IC_{50}$). Spearman's rank correlation coefficient and *p* value are shown, n=18. (**B**) Relationship between pre-CHMI *P. knowlesi* (PvW1) GIA assay data and anti-PvDBPII (PvWI) IgG response measured by ELISA in the purified serum IgG used in the assay. A non-linear regression curve is shown for all samples combined (solid line, $r^2 = 0.58$, n = 18).

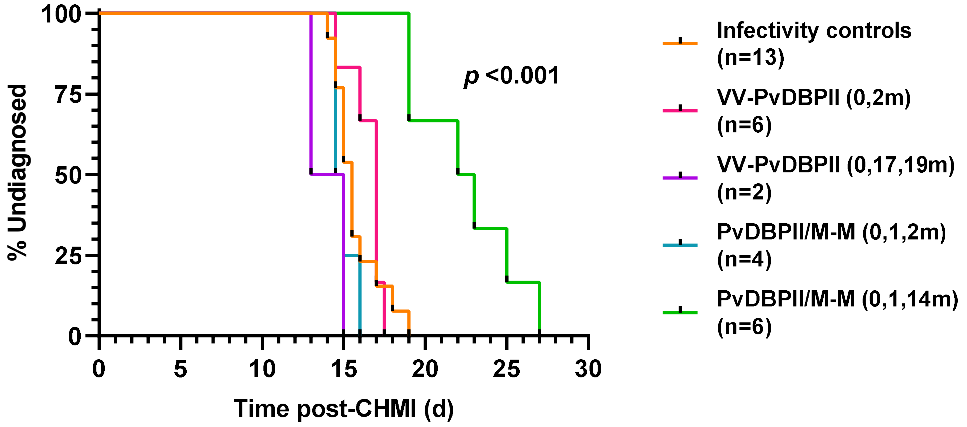

**Figure S6. Kaplan-Meier plot of time to malaria diagnosis.**

Median time to diagnosis was 15.5 days for controls and 22.5 days for PvDBPII/M-M delayed dosing regimen. Pairwise comparison with log-rank test between controls versus vaccine regimen groups was only significant for PvDBPII/M-M (0,1,14m) group ($p$ <0.001).

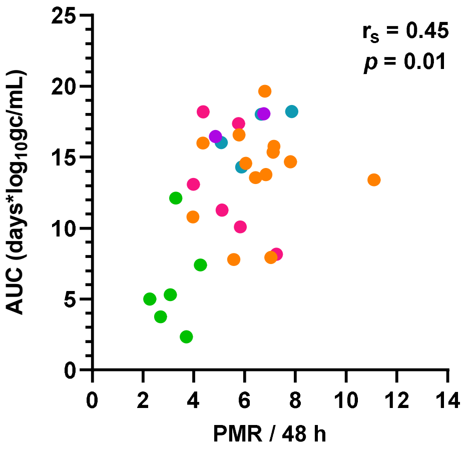

**Figure S7. Relationship between *in vivo* parasite growth as measured by parasite multiplication rate versus $\log_{10}$ cumulative parasitemia.**

Spearman's rank correlation coefficient and $p$ value are shown, n=31.

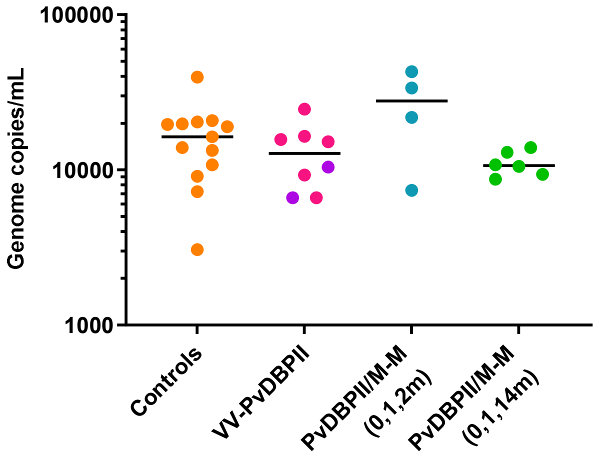

**Figure S8. Malaria qPCR at diagnosis.**

Parasitemia as measured by qPCR in gc/mL just prior to commencing anti-malarial treatment. Individual data and median are shown. No significant differences were observed between the groups as measured by Kruskal-Wallis test with Dunn's multiple comparison post-test.

#### Supplementary Tables

**Table S1. Polymorphisms within region II amino acid sequences of SalI and PvW1 PvDBP.**

| **AA Position** | 261 | 263 | 339 | 340 | 341 | 345 | 353 | 359 | 379 | 430 |
|---|---|---|---|---|---|---|---|---|---|---|
| **PvW1** | L | S | G | K | N | H | T | R | I | L |
| **PvSalI** | F | R | D | E | K | R | S | T | L | - |

Amino acid polymorphisms in region II of PvDBP are shown for those that differ between the *P. vivax* Salvador I strain (vaccine sequence) and the PvW1 clone (used for blood-stage CHMI)[7]. The PvW1 sequence has a leucine insertion between positions 429 and 430 in the SalI sequence.

**Table S2. Baseline demographics of study participants.**

| | | CHMI Controls | VV-PvDBPII | | PvDBPII/M-M | |
|---|---|---|---|---|---|---|
| | | | All vaccinees | CHMI | All vaccinees | CHMI |
| No. of participants | | 13 | 16 | 8 | 16 | 10 |
| Sex no. | Female | 7 | 7 | 4 | 12 | 8 |
| Age - median (range) | | 26 (21, 48) | 25.5 (20, 44) | 29 (21, 41) | 28.5 (19, 44) | 37 (21, 44) |
| Ethnicity no. | White | 10 | 13 | 8 | 15 | 10 |
| | Asian | 1 | 2 | 0 | 0 | 0 |
| | Arab | 1 | 1 | 0 | 0 | 0 |
| | Mixed | 1 | 0 | 0 | 1 | 0 |
| Duffy phenotype no. | Fya+Fyb- | 3 | | 2 | | 1 |
| | Fya-Fyb+ | 2* | | 4 | | 3 |
| | Fya+Fyb+ | 8 | | 2 | | 6 |

VV-PvDBPII = viral-vectored vaccine. PvDBPII/M-M = protein in Matrix-M™ adjuvant vaccine.

* Samples from volunteers who were not heterozygous on Duffy blood group antigen (Fy) serophenotyping were also sent for Duffy blood group antigen genotyping. Only one volunteer (in the control group) with Fya-Fyb+ serophenotype had the Duffy blood group antigen genotype FY*B/FY*B$^{ES}$, whereby the erythrocyte silent (ES) allele has a mutation that prevents Fyb antigen expression in red blood cells.

**Table S3. Laboratory abnormalities following vaccinations.**

| Laboratory abnormality | Number of episodes | | | | | |
|---|---|---|---|---|---|---|
| | ChAd63 PvDBPII | | MVA PvDBPII | PvDBPII/M-M | | |
| | V1 (n=16) | V2 (n=2) | V1 (n=8) | V1 (n=16) | V2 (n=15) | V3 (n=12) |
| Leucopenia | | | | | | |
| Grade 1 | 2 | 0 | 0 | 0 | 0 | 0 |
| Lymphopenia | | | | | | |
| Grade 1 | 5 | 1 | 0 | 0 | 4 | 2 |
| Grade 2 | 6 | 0 | 2 | 1 | 1 | 2 |
| Neutropenia | | | | | | |
| Grade 1 | 3 | 0 | 0 | 0 | 0 | 1 |
| Grade 2 | 2 | 0 | 0 | 0 | 0 | 0 |
| Eosinophilia | | | | | | |
| Grade 1 | 1 | 0 | 0 | 0 | 0 | 0 |
| Hypokalemia | | | | | | |
| Grade 1 | 3 | 1 | 0 | 0 | 0 | 0 |
| Grade 3 | 0 | 1 | 0 | 0 | 0 | 0 |
| Hyperbilirubinemia | | | | | | |
| Grade 1 | 1 | 0 | 0 | 0 | 0 | 0 |
| Anaemia | | | | | | |
| Grade 1 | 0 | 0 | 0 | 0 | 0 | 1 |
| Thrombocytopenia | | | | | | |
| Grade 1 | 0 | 0 | 0 | 1 | 0 | 0 |

Number of episodes of laboratory abnormalities within 28 days following vaccinations with ChAd63 PvDBPII, MVA PvDBPII or PvDBPII/Matrix-M™. Maximal grade of laboratory abnormality, deemed at least possibly related to vaccination, is reported.

**Table S4. Unsolicited adverse events (AE) following ChAd63 or MVA PvDBPII vaccination.**

| Unsolicited AE | Number of episodes of AE | |
|---|---|---|
| | ChAd63 PvDBPII | MVA PvDBPII |
| Gastrointestinal disorders | | |
| Diarrhoea | 1 | 0 |
| Abdominal pain upper | 1 | 0 |
| Vomiting | 1 | 0 |
| Dry mouth | 1 | 0 |
| Respiratory, thoracic and mediastinal disorders | | |
| Rhinitis | 1 | 1 |
| Oropharyngeal pain | 1 | 1 |
| Cough | 0 | 1 |
| Skin and subcutaneous tissue disorders | | |
| Rash | 1 | 0 |
| Musculoskeletal and connective tissue disorders | | |
| Neck pain | 1 | 0 |
| Pain in extremity | 1 | 0 |
| Reproductive system and breast disorders | | |
| Dysmenorrhoea | 2 | 0 |
| Eye disorders | | |
| Dry eye | 1 | 0 |
| General disorders and administration site conditions | | |
| Chest pain | 1 | 0 |
| Nervous system disorders | | |
| Paraesthesia | 0 | 1 |
| Metabolism and nutrition disorders | | |
| Decreased appetite | 0 | 1 |

Unsolicited AEs occurring within 28 days of vaccination and deemed at least possibly related to ChAd63 or MVA PvDBPII vaccination. Number of episodes of unsolicited AE are listed by vaccine and MEDDRA System Organ Class and Preferred Term. Unsolicited AEs were of maximal grade 2 severity.

**Table S5. Unsolicited adverse events (AE) following PvDBPII/M-M vaccination.**

| Unsolicited AE | Number of episodes of AE with PvDBPII/M-M | | | |
|---|---|---|---|---|
| | V1 (n=16) | V2 (n=15) | V3 (n=12) | Total |
| Gastrointestinal disorders | | | | |
| Diarrhoea | 0 | 1 | 0 | 1 |
| General disorders and administration site conditions | | | | |
| Administration site induration | 1 | 0 | 0 | 1 |
| Chest discomfort | 1 | 0 | 0 | 1 |
| Injection site pruritus | 2 | 0 | 0 | 2 |
| Injection site swelling | 1 | 0 | 0 | 1 |
| Swelling | 1 | 0 | 0 | 1 |
| Infections and infestations | | | | |
| Rhinitis | 1 | 0 | 0 | 1 |
| Musculoskeletal and connective tissue disorders | | | | |
| Back pain | 2 | 1 | 0 | 3 |
| Pain in extremity | 2 | 1 | 0 | 3 |
| Nervous system disorders | | | | |
| Dizziness | 1 | 0 | 0 | 1 |
| Headache | 0 | 2 | 1 | 3 |
| Migraine | 1 | 0 | 0 | 1 |
| Hypoaesthesia | 0 | 1 | 0 | 1 |
| Paraesthesia | 2 | 0 | 0 | 2 |
| Somnolence | 0 | 1 | 0 | 1 |
| Taste disorder | 1 | 0 | 0 | 1 |
| Psychiatric disorders | | | | |
| Euphoric mood | 1 | 0 | 0 | 1 |
| Insomnia | 1 | 1 | 1 | 3 |
| Tearfulness | 1 | 0 | 0 | 1 |
| Reproductive system and breast disorders | | | | |
| Dysmenorrhoea | 0 | 1 | 0 | 1 |
| Respiratory, thoracic and mediastinal disorders | | | | |
| Nasal congestion | 1 | 0 | 0 | 1 |
| Oropharyngeal pain | 0 | 1 | 0 | 1 |
| Throat irritation | 1 | 1 | 0 | 2 |

Unsolicited AEs occurring within 28 days of vaccination and deemed at least possibly related to PvDBPII/M-M vaccination. Number of episodes of unsolicited AE following the first, second and third vaccination are listed by MEDDRA System Organ Class and Preferred Term. Unsolicited AEs were of maximal grade 2 severity.

**Table S6. Summary of parasite multiplication rate (PMR) analysis**

| | Controls | VV-PvDBPII | PvDBPII/M-M (0,1,2m) | PvDBPII/M-M (0,1,14m) |
|---|---|---|---|---|
| No of volunteers | 13 | 8 | 4 | 6 |
| Median PMR per 48 h | 6.8 | 5.4 | 6.3 | 3.2 |
| Range PMR per 48 h | 4.0 to 11.1 | 4.0 to 7.3 | 5.1 to 7.9 | 2.3 to 4.3 |
| D'Agostino & Pearson $K^2$ test | $p = 0.02$ | $p = 0.78$ | $p = 0.57$ | |
| Mann-Whitney test* | | $p = 0.14$ | $p = 0.01$ | |

*Two tailed *p* value reported for Mann-Whitney test comparing controls with each vaccine group.

**Table S7. Analysis of PMR by study group and Duffy blood group serophenotype**

| | Univariate predictor | | | Adjusted for other variable | | |
|---|---|---|---|---|---|---|
| Variable | Estimate | 95% CI | *p* value | Estimate | 95% CI | *p* value |
| Intercept | 6.6 | 5.8 to 7.4 | <0.001 | 6.3 | 5.5 to 7.2 | <0.001 |
| Study group | | | | | | |
| Controls | 0 | | | 0 | | |
| VV-PvDBPII | -1.1 | -2.4 to 0.2 | 0.08 | -1.2 | -2.5 to 0.2 | 0.08 |
| PvDBPII/M-M (0,1,2m) | -0.2 | -1.9 to 1.4 | 0.76 | 0.04 | -1.6 to 1.7 | 0.96 |
| PvDBPII/M-M (0,1,14m) | -3.4 | -4.8 to -2.0 | <0.001 | -3.3 | -4.7 to -1.9 | <0.001 |
| DARC serophenotype | | | | | | |
| Fya+b+ | | | | 0 | | |
| Fya+b- | | | | 1.2 | -0.1 to 2.6 | 0.08 |
| Fya-b+ | | | | 0.02 | -1.2 to 1.3 | 0.97 |
| R squared | 0.49 | | | 0.56 | | |
| No observations | 31 | | | 31 | | |

Multiple linear regression was used to test if PMR differed significantly between different Duffy blood group antigen (DARC) serophenotypes after controlling for vaccination group.

**Table S8. Malaria qPCR data (gc/mL) for CHMI in September 2019.**

| Trial | Group | DoD | D7 | D8 | D9 | D10 | D11 | D11.5 | D12 | D12.5 | D13 | D13.5 | D14 | D14.5 | D15 | D15.5 | D16 | D16.5 | D17 | D17.5 |
|---|---|---|---|---|---|---|---|---|---|---|---|---|---|---|---|---|---|---|---|---|
| VAC069 | 6 | 15.5 | N | N | 45 | 104 | 209 | | 270 | | 1379 | 1391 | 2244 | 3670 | 8921 | 9574 | 16345 | | | |
| VAC069 | 6 | 14.5 | N | 9 | 72 | N | 265 | | 778 | | 3365 | 2835 | 4283 | 17392 | 19589 | | | | | |
| VAC071 | 1 | 17 | 11 | 9 | N | 16 | 54 | | 198 | | 608 | | 792 | | 3098 | 3281 | 3694 | 4831 | 16135 | 15663 |
| VAC071 | 1 | 17 | N | N | 26 | 13 | 80 | | 94 | | 362 | | 652 | | 2768 | 2751 | 4192 | 3832 | 15176 | |
| VAC071 | 1 | 17.5 | N | 27 | 31 | 44 | 74 | | 123 | | 527 | | 1285 | 889 | 2745 | 2295 | 2465 | 2914 | 9588 | 9263 |

Malaria qPCR data used in PMR modelling. Top row represents day (D) of follow-up visit post blood-stage CHMI. DoD = timepoint at which malaria diagnostic criteria were reached. VAC069 Group 6 = infectivity controls; VAC071 Group 1 = VV-PvDBPII given at 0, 2 months. Treatment in some volunteers was started half a day after reaching malaria diagnostic criteria. qPCR data shown for samples taken prior to starting treatment. qPCR negative values for all three triplicate readings in the assay are indicated by ‘N’. Squares highlighted in grey indicate negative or < 20 gc/mL which is below minimum positive reporting criteria and these datapoints were removed for PMR modelling. Datapoints which would not have been taken during CHMI in October 2021 due to changes in protocol were removed for PMR modelling and are not shown.

**Table S9. Malaria qPCR data (gc/mL) for CHMI in May 2021.**

| Trial | Group | DoD | D7 | D8 | D9 | D10 | D11 | D11.5 | D12 | D12.5 | D13 | D13.5 | D14 | D14.5 | D15 | D15.5 | D16 | D16.5 | D17 | D17.5 | D18 |
|---|---|---|---|---|---|---|---|---|---|---|---|---|---|---|---|---|---|---|---|---|---|
| VAC069 | 9 | 15.5 | N | N | 74 | 220 | 316 | | 408 | | 1678 | 1315 | 3625 | 6183 | 8674 | 9095 | | | | | |
| VAC069 | 9 | 18 | 10 | N | 8 | N | 125 | | 88 | | 291 | | 814 | 1252 | 1652 | 1805 | 2152 | 6550 | 7261 | 6161 | 12050 |
| VAC069 | 9 | 19 | N | N | 26 | 37 | 98 | | 62 | | 268 | | 298 | | 1111 | 1262 | 1617 | 2121 | 6005 | 6395 | 4758 |
| VAC069 | 9 | 14 | 32 | 35 | 108 | 182 | 1059 | 748 | 1121 | 2572 | 8025 | 7805 | 14766 | 18983 | | | | | | | |
| VAC069 | 9 | 16 | 11 | N | 47 | 74 | 311 | | 298 | | 1669 | 1544 | 3316 | 6509 | 8709 | 9292 | 15568 | 39496 | | | |
| VAC069 | 9 | 14.5 | N | N | 50 | 96 | 370 | | 754 | | 2641 | 2335 | 4847 | 12160 | 7238 | | | | | | |
| VAC069 | 9 | 15 | N | N | 113 | 183 | 485 | | 556 | | 3001 | 2476 | 5926 | 7904 | 16068 | 20776 | | | | | |
| VAC079 | 1 | 19 | 6 | 21 | 17 | 31 | 79 | | 73 | | 316 | | 379 | | 798 | | 1038 | 1474 | 3570 | 3555 | 3665 |
| VAC079 | 1 | 19 | N | N | 18 | 15 | 77 | | 135 | | 155 | | 442 | | 719 | | 1101 | 1578 | 3775 | 3861 | 2797 |
| VAC079 | 1 | 25 | N | N | N | 15 | 6 | | 25 | | 34 | | 52 | | 70 | | 166 | | 429 | | 306 |
| VAC079 | 1 | 22 | N | N | N | N | 5 | | 18 | | 23 | | 100 | | 60 | | 243 | | 568 | | 793 |
| VAC079 | 1 | 27 | 1 | N | N | N | 27 | | 23 | | 34 | | 28 | | 27 | | 107 | | 49 | | 57 |
| VAC079 | 1 | 23 | N | N | 7 | N | 27 | | 6 | | 40 | | 40 | | 151 | | 194 | | 370 | | 494 |

| Trial | Group | DoD | D18.5 | D19 | D19.5 | D20 | D20.5 | D21 | D21.5 | D22 | D22.5 | D23 | D23.5 | D24 | D24.5 | D25 | D25.5 | D26 | D26.5 | D27 | D27.5 |
|---|---|---|---|---|---|---|---|---|---|---|---|---|---|---|---|---|---|---|---|---|---|
| VAC069 | 9 | 15.5 | | | | | | | | | | | | | | | | | | | |
| VAC069 | 9 | 18 | 20348 | | | | | | | | | | | | | | | | | | |
| VAC069 | 9 | 19 | 9432 | 19781 | | | | | | | | | | | | | | | | | |
| VAC069 | 9 | 14 | | | | | | | | | | | | | | | | | | | |
| VAC069 | 9 | 16 | | | | | | | | | | | | | | | | | | | |
| VAC069 | 9 | 14.5 | | | | | | | | | | | | | | | | | | | |
| VAC069 | 9 | 15 | | | | | | | | | | | | | | | | | | | |
| VAC079 | 1 | 19 | 3769 | 13066 | 10752 | | | | | | | | | | | | | | | | |
| VAC079 | 1 | 19 | 6564 | 12971 | | | | | | | | | | | | | | | | | |
| VAC079 | 1 | 25 | | 968 | | 745 | | 2043 | 2019 | 3302 | 2464 | 4198 | 4425 | 6010 | 5591 | 15486 | 10533 | | | | |
| VAC079 | 1 | 22 | | 2504 | 2008 | 3138 | 3749 | 8451 | 7364 | 12330 | 9359 | | | | | | | | | | |
| VAC079 | 1 | 27 | | 86 | | 173 | | 397 | | 565 | | 930 | | 1597 | | 3766 | 3290 | 5621 | 5404 | 12220 | 8699 |
| VAC079 | 1 | 23 | | 1242 | | 1338 | | 3417 | | 4662 | 7714 | 13665 | 13903 | | | | | | | | |

Malaria qPCR data used in PMR modelling. Top row represents day (D) of follow-up visit post blood-stage CHMI. DoD = timepoint at which malaria diagnostic criteria were reached. VAC069 Group 9 = infectivity controls; VAC079 Group 1 = PvDBPII/M-M given at 0, 1, 14 months. Treatment in some volunteers was started half a day after reaching malaria diagnostic criteria. qPCR data shown for samples taken prior to starting treatment. qPCR negative values for all three triplicate readings in the assay are indicated by 'N'. Squares highlighted in grey indicate negative or < 20 gc/mL which is below minimum positive reporting criteria and these datapoints were removed for PMR modelling. Datapoints which would not have been taken during CHMI in October 2021 due to changes in protocol were removed for PMR modelling and are not shown.

**Table S10. Malaria qPCR data (gc/mL) for CHMI in October 2021.**

| Trial | Group | DoD | D7 | D8 | D9 | D10 | D11 | D11.5 | D12 | D12.5 | D13 | D13.5 | D14 | D14.5 | D15 | D15.5 | D16 | D16.5 | D17 | D17.5 |
|---|---|---|---|---|---|---|---|---|---|---|---|---|---|---|---|---|---|---|---|---|
| VAC069 | 12 | 15 | **8** | **28** | **66** | **124** | **604** | | **806** | | **2823** | **2487** | **3759** | **3551** | **14373** | **10759** | | | | |
| VAC069 | 12 | 15 | **17** | **17** | **71** | **119** | **1021** | **804** | **1617** | **1727** | **3405** | **4145** | **5742** | **6828** | **17155** | **13910** | | | | |
| VAC069 | 12 | 17 | **N** | **9** | **N** | **7** | **73** | | **46** | | **510** | | **501** | | **1735** | **1792** | **3600** | **4258** | **12302** | **13366** |
| VAC069 | 12 | 15.5 | **N** | **61** | **53** | **73** | **401** | | **767** | | **1552** | **1096** | **2597** | **2245** | **5737** | **3057** | | | | |
| VAC071 | 2 | 13 | **47** | **54** | **128** | **279** | **1252** | **1196** | **2498** | **2221** | **9116** | **10400** | | | | | | | | |
| VAC071 | 2 | 15 | **N** | **28** | **81** | **162** | **447** | | **950** | | **1941** | **1523** | **2321** | **1996** | **6614** | | | | | |
| VAC071 | 3 | 16 | **27** | **60** | **73** | **107** | **620** | | **912** | | **1918** | **2059** | **2644** | **6468** | **7675** | **4376** | **12306** | **24606** | | |
| VAC071 | 3 | 14.5 | **N** | **70** | **76** | **219** | **517** | | **885** | | **3010** | **3061** | **5275** | **6615** | | | | | | |
| VAC071 | 3 | 17 | **9** | **N** | **17** | **39** | **123** | | **102** | | **400** | | **787** | | **2660** | **1875** | **4213** | **5979** | **15424** | **16420** |
| VAC079 | 2 | 14.5 | **17** | **20** | **169** | **242** | **1227** | **989** | **1517** | **2934** | **5399** | **6277** | **8780** | **17594** | **33726** | | | | | |
| VAC079 | 2 | 14.5 | **N** | **107** | **129** | **90** | **1244** | **852** | **1657** | **1751** | **2228** | **5138** | **8719** | **11292** | **42914** | | | | | |
| VAC079 | 2 | 15 | **9** | **14** | **61** | **46** | **536** | | **652** | | **1961** | **2072** | **2070** | **1030** | **10301** | **7374** | | | | |
| VAC079 | 2 | 16 | **8** | **21** | **130** | **65** | **503** | | **611** | | **1855** | **1765** | **2681** | **4131** | **7432** | **5408** | **13620** | **21801** | | |

Malaria qPCR data used in PMR modelling. Top row represents day (D) of follow-up visit post blood-stage CHMI. DoD = timepoint at which malaria diagnostic criteria was reached. VAC069 Group 12 = infectivity controls; VAC071 Group 2 = VV-PvDBPII given at 0, 17, 19 months; VAC079 Group 2 = PvDBPII/M-M given at 0, 1, 2 months. Treatment in some volunteers was started half a day after reaching malaria diagnostic criteria. qPCR data shown for samples taken prior to starting treatment. qPCR negative values for all three triplicate readings in the assay are indicated by 'N'. Squares highlighted in grey indicate negative or < 20 gc/mL which is below minimum positive reporting criteria and these datapoints were removed for PMR modelling.

**Table S11. PvDBPII peptides used for T cell stimulation.**

| Peptide Number | N-terminus | Amino Acid Sequence | C-terminus |
|---|---|---|---|
| 1 | H- | DHKKTISSAIINHAFLQNTVGSG(261) | -NH2 |
| 2 | Biotin- | SGSGAIINHAFLQNTVMKNCNYKR | -NH2 |
| 3 | Biotin- | SGSGQNTVMKNCNYKRKRRERDWD | -NH2 |
| 4 | Biotin- | SGSGNYKRKRRERDWDCNTKKDVC | -NH2 |
| 5 | Biotin- | SGSGRDWDCNTKKDVCIPDRRYQL | -NH2 |
| 6 | Biotin- | SGSGKDVCIPDRRYQLCMKELTNL | -NH2 |
| 7 | Biotin- | SGSGRYQLCMKELTNLVNNTDTNF | -NH2 |
| 8 | Biotin- | SGSGLTNLVNNTDTNFHRDITFRK | -NH2 |
| 9 | Biotin- | SGSGDTNFHRDITFRKLYLKRKLI | -NH2 |
| 10 | Biotin- | SGSGTFRKLYLKRKLIYDAAVEGD | -NH2 |
| 11 | Biotin- | SGSGRKLIYDAAVEGDLLLKLNNY | -NH2 |
| 12 | Biotin- | SGSGVEGDLLLKLNNYRYNKDFCK | -NH2 |
| 13 | Biotin- | SGSGLNNYRYNKDFCKDIRWSLGD | -NH2 |
| 14 | Biotin- | SGSGDFCKDIRWSLGDFGDIIMGT | -NH2 |
| 15 | Biotin- | SGSGSLGDFGDIIMGTDMEGIGYS | -NH2 |
| 16 | Biotin- | SGSGIMGTDMEGIGYSKVVENNLR | -NH2 |
| 17 | Biotin- | SGSGIGYSKVVENNLRSIFGTDEK | -NH2 |
| 18 | Biotin- | SGSGNNLRSIFGTDEKAQQRRKQW | -NH2 |
| 19 | Biotin- | SGSGTDEKAQQRRKQWWNESKAQI | -NH2 |
| 20 | Biotin- | SGSGRKQWWNESKAQIWTAMMYSV | -NH2 |
| 21 | Biotin- | SGSGKAQIWTAMMYSVKKRLKGNF | -NH2 |
| 22 | Biotin- | SGSGMYSVKKRLKGNFIWICKLNV | -NH2 |
| 23 | Biotin- | SGSGKGNFIWICKLNVAVNIEPQI | -NH2 |
| 24 | Biotin- | SGSGKLNVAVNIEPQIYRWIREWG | -NH2 |
| 25 | Biotin- | SGSGEPQIYRWIREWGRDYVSELP | -NH2 |
| 26 | Biotin- | SGSGREWGRDYVSELPTEVQKLKE | -NH2 |
| 27 | Biotin- | SGSGSELPTEVQKLKEKCDGKINY | -NH2 |
| 28 | Biotin- | SGSGKLKEKCDGKINYTDKKVCKV | -NH2 |
| 29 | Biotin- | SGSGKINYTDKKVCKVPPCQNACK | -NH2 |
| 30 | Biotin- | SGSGVCKVPPCQNACKSYDQWITR | -NH2 |
| 31 | Biotin- | SGSGNACKSYDQWITRKKNQWDVL | -NH2 |
| 32 | Biotin- | SGSGWITRKKNQWDVLSNKFISVK | -NH2 |
| 33 | Biotin- | SGSGWDVLSNKFISVKNAEKVQTA | -NH2 |
| 34 | Biotin- | SGSGISVKNAEKVQTAGIVTPYDI | -NH2 |
| 35 | Biotin- | SGSGVQTAGIVTPYDILKQELDEF | -NH2 |
| 36 | Biotin- | SGSGPYDILKQELDEFNEVAFENE | -NH2 |
| 37 | Biotin- | SGSGLDEFNEVAFENEINKRDGAY | -NH2 |
| 38 | Biotin- | SGSGFENEINKRDGAYIELCVCSV | -NH2 |
| 39 | Biotin- | SGSGDGAYIELCVCSVEEAKKNTQ | -NH2 |
| 40 | Biotin- | SGSGIELCVCSVEEAKKNTQEVVT | -OH |

The PvDBPII SalI amino acid sequence was used to design 20mer peptides overlapping by 12 amino acids and these were synthesized by Mimotopes, Australia. Each stock was reconstituted to 50 mg/mL in DMSO. A 200 µg/peptide/mL working stock of PvDBPII peptides was prepared by adding an equal amount of each peptide to cell culture medium for a final total peptide concentration of 8 mg/mL.
